# Early life predictors of late life cerebral small vessel disease in four prospective cohort studies

**DOI:** 10.1101/2021.09.13.21252900

**Authors:** Ellen V Backhouse, Susan D Shenkin, Andrew M McIntosh, Mark E Bastin, Heather C Whalley, Maria Valdez Hernandez, Susana Muñoz Maniega, Mat Harris, Aleks Stolicyn, Archie Campbell, Douglas Steele, Gordon D Waiter, Anca-Larisa Sandu, Jennifer MJ Waymont, Alison D Murray, Simon R Cox, Susanne R. de Rooij, Tessa J. Roseboom, Joanna M Wardlaw

## Abstract

Development of cerebral small vessel disease, a major cause of stroke and dementia, may be influenced by early life factors. It is unclear whether these relationships are independent of each other, of adult socioeconomic status or of vascular risk factor exposures.

We examined associations between factors from birth (ponderal index, birth weight), childhood (IQ, education, socioeconomic status), adult small vessel disease, and brain volumes, using data from four prospective cohort studies: STratifying Resilience And Depression Longitudinally (STRADL) (*n*=1080; mean age=59 years); The Dutch Famine Birth cohort (*n*=118; mean age=68 years); the Lothian Birth Cohort 1936 (LBC1936; *n*=617; mean age=73 years), and the Simpson’s cohort (*n*=110; mean age=78 years). We analysed each small vessel disease feature individually and summed to give a total small vessel disease score (range 1-4) in each cohort separately, then in meta-analysis, adjusted for vascular risk factors and adult socioeconomic status.

Higher birth weight was associated with fewer lacunes (*OR* per 100g, 0.93 *95%CI*=0.88-0.99), fewer infarcts (*OR*=0.94 *95%CI*=0.89-0.99), and fewer perivascular spaces (*OR*=0.95 *95%CI*=0.91-0.99). Higher childhood IQ was associated with lower white matter hyperintensity burden (*OR* per IQ point=0.99 95%CI 0.98-0.998), fewer infarcts (OR=0.98, 95%CI=0.97-0.998), fewer lacunes (*OR*=0.98, *95%CI*=0.97-0.999), and lower total small vessel disease burden (*OR*=0.98, *95%CI*=0.96-0.999). Low education was associated with more microbleeds (*OR*=1.90 *95%CI*=1.33-2.72) and lower total brain volume (*MD*=-178.86cm^3^, *95%CI*=-325.07- -32.66). Low childhood socioeconomic status was associated with fewer lacunes (*OR*=0.62, *95%CI*=0.40-0.95).

Early life factors are associated with worse small vessel disease in later life, independent of each other, vascular risk factors and adult socioeconomic status. Risk for small vessel disease may originate in early life and provide a mechanistic link between early life factors and risk of stroke and dementia. Policies investing in early child development may contribute to improve lifelong brain health to prevent dementia and stroke in older age.

## Introduction

Cerebral small vessel disease (SVD) is common at older ages^1^ and causes 20-25% of strokes and up to 45% of dementias, either as vascular or mixed with Alzheimer’s disease.^2^ It is responsible for up to a fifth of all strokes, doubles the risk of future stroke and worsens post-stroke recovery.^3^ SVD is detected on neuroimaging or post mortem^4^ as white matter hyperintensities (WMH), lacunes, microbleeds, perivascular spaces (PVS), acute lacunar infarcts and brain atrophy.^4,5^ Several demographic and clinical factors are associated with increased risk of SVD, including adult socioeconomic status (SES), hypertension and smoking.^6,7^ However, a large proportion of the variance in the presence and severity of SVD is unexplained by vascular risk factors^7^ and factors from earlier in life may also be important.^8^

The Developmental Origins of Adult Heath and Disease (DOHAD) hypothesis^9^ proposes that adverse environmental exposures occurring during gestation can cause permanent changes in fetal development resulting in increased vulnerability to chronic diseases in adulthood. Factors affecting foetal growth such as stress and poor nutrition^10,11^ are often hard to measure but anthropometric measures such as birth weight and ponderal index (birth weight/birth length^3^) can be used as proxy measures.^12^ Additional confounding or mediating factors in childhood may also affect later disease risk.^13^ A recent meta-analysis^14^ found that lower levels of childhood IQ, poorer childhood SES, and less education increased the risk of SVD in later life by approximately 17-39%. However, it is not clear if these relationships are independent of each other, or if they persist after adjustment for vascular risk factors and adult SES. Few studies have examined the effect of these early life factors in combination and many rely on childhood measures assessed retrospectively in adulthood so may be subject to recall bias.

We examined the relationships between birth and childhood factors and total and individual components of SVD and brain volumes, after adjustment for each other and common adult risk factors, in four well-phenotyped prospective cohort studies: STratifying Resilience and Depression Longitudinally (STRADL).^15^ the Dutch Famine Birth Cohort,^16^ the Lothian Birth Cohort 1936 (LBC1936),^17^ and the Simpson’s cohort.^18^ All had information on education and SES, and three cohorts had IQ measured during childhood. All underwent brain imaging between the ages of 59 and 85 years. We hypothesised that low birth weight, low childhood IQ, low education and low childhood SES would be associated with increased SVD, independent of each other, vascular risk factors and adult SES.

## Materials and methods

### Participants

The recruitment procedures and inclusion criteria for STRADL,^15^ the Dutch Famine Birth Cohort,^16^ the LBC1936,^17^ and the Simpson’s cohort^18^ have been described previously in detail (see Supplementary Fig. 1a-d for recruitment flow charts). All subjects were community dwelling.

### STRADL

STRADL is a population based study of 1,198 adults recruited from the Generation Scotland: Scottish Family Health Study (GS:SFHS) and two Scottish longitudinal birth cohorts, the Aberdeen Children of the 1950s (ACONF) cohort^19^ and the Walker cohort.^20^ ACONF consists of surviving participants of the Aberdeen Child Development Survey (ACDS), a population based study of schoolchildren in Aberdeen, conducted in 1962-64. The Walker cohort is a database of over 48,000 birth records of babies born in hospital in Dundee, between 1952 and 1966. In 2015 eligible participants were sent postal questionnaires and between 2015 and 2019 1,188 attended in person assessments. MRI and childhood data were available for 1080 participants (ACONF 268; Walker 201; GS:SFHS 611) (40% female; mean age= 59.3 years, *SD*=10.1).

### The Dutch Famine Birth cohort

The Dutch Famine Birth Cohort consists of 2,414 individuals born in the Wilhelmina Gasthuis hospital in Amsterdam between 1^st^ November 1943 and 28^th^ February 1947, a proportion of whom were exposed to the Dutch famine of 1944-1945 in utero. 151 surviving cohort members were recruited for an MRI study in 2012 of which 118 had MRI and childhood data (56% female; mean age=67.5 years, *SD*=0.9).

### The Lothian Birth cohort 1936 (LBC1936)

The LBC1936 consists of 1,091 community dwelling adults born in 1936 and living in the Lothian area of Scotland. All are surviving participants of the Scottish Mental Health Survey 1947 which was a cognitive ability test administered to all age 11 school children in Scotland in 1947. Between 2007 and 2009 680 of the original 1091 cohort members underwent MRI, all with childhood data (47% female; mean age 72.7 years, *SD*=0.7).

### The Simpson’s Cohort

The Simpson’s cohort consists of 130 individuals born 1921-1926 in three Edinburgh hospitals. In 2000 28 people were recruited as part of the Lothian Birth Cohort 1921, 19 were traced through hospital records from 1921 and 80 people were recruited through local advertisements. MRI and childhood data were available for 110 people (67% female, mean age= 78.4 years, *SD*=1.5).

Participants in all cohorts provided written informed consent and research was approved by Local or Multicentre Research Ethics Committees. (STRADL:14/SS/0039; LBC1936: MREC/01/0/56 and LREC/2003/2/29; Simpson’s cohort LREC 1702/1998/4/183– Amendment).

### Early life factors

The early life data available varied between cohorts (Fig. 1). Where possible, data were harmonised to allow direct comparison between the studies. We examined birth weight in grams (all cohorts) and ponderal index (birth weight/birth length^3^) (Dutch Famine Birth Cohort, LBC1936 and Simpson’s cohort). In childhood, we examined: childhood IQ (STRADL, LBC1936 and Simpson’s cohort) measured using raw test scores adjusted for age at testing and placed on an IQ scale; education (all cohorts) dichotomised at compulsory education (STRADL), lower secondary (Dutch Famine cohort) and 11 years (LBC1936 and Simpson’s cohort); and childhood SES (all cohorts) classified according to parental occupation (manual and non-manual). Further details are provided in Supplementary Table 1.

**Figure 1:**
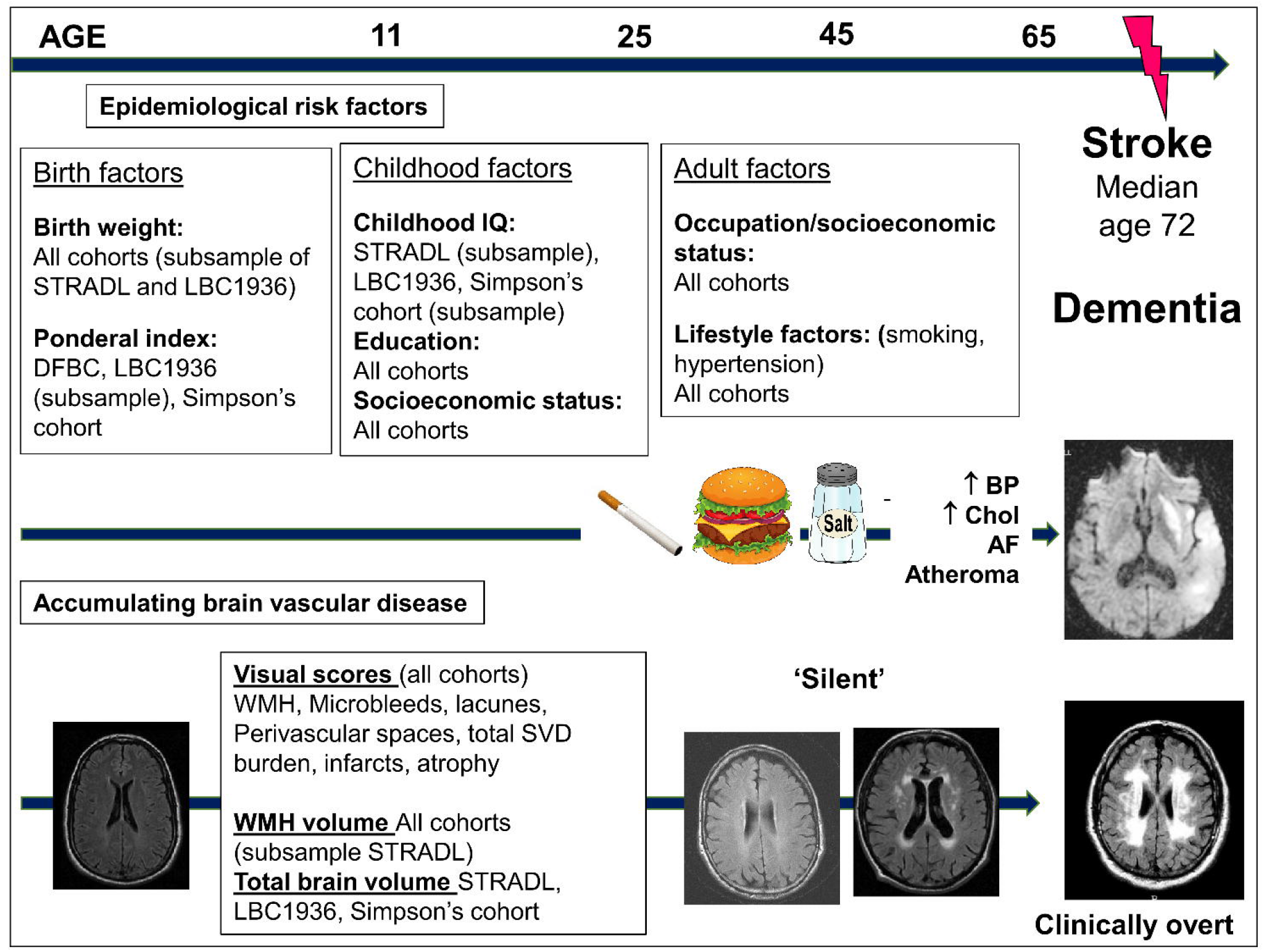
The lifecourse perspective of the risk of SVD and stroke. Adapted from Figure 1, Backhouse *Curr Epidemiol Rep* 2015 2:172-179.

### MRI acquisition and analysis

Brain imaging acquisition for STRADL,^21^ the Dutch Famine birth cohort,^22^ LBC1936^23^ and the Simpson’s cohort^24^ have been described previously. Participants were scanned on a Philips Achieva 3.0T TX (STRADL, Aberdeen), Siemens 3T Prisma-FIT (STRADL Dundee), a 3T Philips Ingenia (Best, the Netherlands) with a 16-channel DStream Head-Spin coil (Dutch Famine cohort), or the same 1.5T GE Signa scanner operating in research mode in its original LX format (Simpson’s cohort) or following an upgrade to HDx format (LBC1936) (Supplementary Table 2).

### SVD visual ratings

Trained researchers using the same rating methods and, blind to all other data, performed all image analyses. An experienced certified and registered neuroradiologist (JMW) crosschecked 20% of scans. Presence of WMH, lacunes, micro-bleeds and perivascular spaces were rated according to STRIVE criteria and established protocols, published previously using validated visual scales,^23,25,26^ converted to dichotomous point scores and summed to create the total SVD score (0-4; higher score represents higher SVD burden).^6,27-29^ We noted any imaging evidence of infarcts in the cortical or subcortical regions using a validated stroke lesion rating scale.^30^ Superficial and deep atrophy scores were coded separately using a valid template,^31^ summed to give a total score and dichotomised into ‘none or mild’ and ‘moderate or severe’.

### WMH volumes and whole brain volume

We conducted structural image analysis, blind to all non-imaging data, including measurements of volumes of the intracranial compartment (ICV), whole brain and total WMH volume in STRADL, LBC1936 and the Simpson’s cohort and WMH volume only in the Dutch Famine Birth cohort. For tissue segmentation we used the processing protocol with the Lesion Growth Algorithm (LGA), provided by the Lesion Segmentation Toolbox for SPM (STRADL) and a semiautomatic segmentation tool MCMxxxVI previously validated^32^ (LBC1936 and Simpson’s cohort). We visually inspected all segmented images and manually edited any incorrectly classified tissues. Analyses were performed using Freesurfer 5.3 and Analyze^™^ software.

### Statistical analysis

We assessed descriptive characteristics using means, standard deviations (SD), medians and interquartile range (IQR), counts and percentages as appropriate. We used *χ*^2^ for categorical data and Mann-Whitney U-Test for continuous data to compare differences between participants who underwent MRI and those who did not and to examine gender differences in SVD burden.

Few in the cohorts had the highest SVD scores, which likely reflects the generally good health of the cohorts. We therefore dichotomised the SVD score into 0-1 (“no or mild disease”) and 2-4 (“moderate-severe disease”).

We performed logistic regression for differences in early life factors for higher vs lower SVD scores and for presence of each individual SVD component and linear regression analysis to assess early life factors and brain volumes. Brain volumes were adjusted for intracranial volume (ICV). For all main analyses we analysed the cohorts individually and meta-analysed them using a random effects model in Review Manager 5.3. Due to the small sample size for some analyses we did not adjust for all available vascular risk factors. Based on previous research^6,7^ we included age, sex, hypertension, smoking behaviour and adult SES at the time of the MRI (manual vs non-manual occupation) as covariates in all models. We adjusted analyses including birth weight and ponderal index for gestational age taken from birth records. We performed further multiple regression analyses adjusting for the other early life factors and where sample size allowed, using an event per variable of 10, vascular risk factors and SES in adulthood. A Bonferroni correction for multiple testing was not appropriate, as the variables are not independent. Therefore to mitigate the problem of multiple testing, we defined our hypotheses a priori based on our previous meta-analysis.^33^

All analyses were performed using SPSS version 24 (IBM Corp., Armonk, NY) using pairwise deletion to deal with missing data.

### Data availability

The data that support the findings of this study are available upon reasonable request.

## Results

Demographic and key characteristics of all participants are displayed in Table 1A-C.

**Table 1.** **A-C:** Demographic and health characteristics (A), early life characteristics (B) and imaging characteristics (C) of STRADL, the Dutch Famine Birth cohort, the LBC1936 and the Simpson’s cohort.

**(A) Demographic and health characteristics**
|  | STRADL |  | Dutch Famine |  | LBC1936 |  | Simpson's |  |
| --- | --- | --- | --- | --- | --- | --- | --- | --- |
|  | Total<br>n | n (%) | Total<br>n | n (%) | Total<br>n | n (%) | Total<br>n | n (%) |
| Age (y) at MRI, mean (SD),<br>range | 1080 | 59.3 (10.1),<br>26-84 | 118 | 67.5 (0.9),<br>65-69 | 685 | 72.7 (0.7),<br>71-74 | 110 | 78.4 (1.5),<br>75-81 |
| Sex, male | 1080 | 437 (40.5) | 118 | 52 (44.1) | 685 | 361 (52.7) | 110 | 33 (30) |
| Manual adult SES | 1070 | 345 (31.9) | 118 | 44 (37.3) | 674 | 141 (20.9) | 110 | 64 (58.2) |
| **Health history** |  |  |  |  |  |  |  |  |
| History of stroke | 1080 | 33 (3.1) | 117 | 3 (2.6) | 685 | 47 (6.9) | 110 | 16 (14.6) |
| Hypertension | 1080 | 299 (27.7) | 117 | 62 (53.0) | 685 | 336 (49.1) | 110 | 49 (44.6) |
| Diabetes | 1069 | 84 (7.9) | 118 | 24 (20.3) | 685 | 72 (10.5) | 110 | 7 (6.4) |
| Hypercholesterolemia | 930 | 221 (23.7) | 117 | 56 (47.9) | 685 | 287 (41.9) | - | - |
| Smoking history | 962 |  | 118 |  | 685 |  | 110 |  |
| Ever smoker |  | 435 (40.3) |  | 72 (61.0) |  | 362 (52.9) |  | 60 (54.6) |
| Never smoked |  | 527 (48.8) |  | 46 (39.0) |  | 323 (47.2) |  | 50 (45.5) |

SES: socioeconomic status; - is used where data not available;

**(B) Early life characteristics**
| Variable | STRADL |  | Dutch Famine |  | LBC1936 |  | Simpson's |  |
| --- | --- | --- | --- | --- | --- | --- | --- | --- |
|  | Total<br>N | n (%) | Total<br>N | n (%) | Total<br>N | n (%) | Total<br>N | n (%) |
| Years of birth | 1933-1993<br>Median year 1955 |  | 1944-1947 |  | 1936 |  | 1921-1926 |  |
| Birth factors |  |  |  |  |  |  |  |  |
| Ponderal index (kg/m<sup>3</sup>)<br>mean (SD) | - | - | 115 | 26.2 (2.3) | 79 | 27.3 (5.3) | 107 | 25.8 (4.2) |
| Birth weight (g), mean<br>(SD) | 154 | 3309.3 (529.4) | 118 | 3417.5 (503.4) | 140 | 3351.5 (482.1) | 110 | 3333.6 (457.2) |
| Low birth weight<br>($<5$lbs), n (%) | 253 | 6 (2.4) | - | - | - | - | - | - |
| Birth length (cm), mean<br>(SD) | - | - | 118 | 51.9 (8.0) | 79 | 50.0 (3.3) | 107 | 50.7 (2.8) |
| Childhood factors |  |  |  |  |  |  |  |  |
| Childhood IQ | 246 | 102.0 (8.9) | - | - | 648 | 100.8 (15.3) | 30 | 101.7 (14.5) |
| Low vs high level of<br>education<sup>a</sup> | 1078 | 259 (24.0) | 118 | 74 (62.7) | 685 | 491 (71.7) | 110 | 89 (80.9) |
| Manual Father's<br>occupation | 1070 | 719 (67.2) | 96 | 64 (66.7) | 627 | 465 (74.2) | 110 | 76 (69.1) |

<sup>a</sup>Low education defined as compulsory education and below (STRADL), lower secondary school and below (Dutch Famine Birth cohort) and 11 years and below (LBC 1936 and Simpson's cohort).

**(C) Imaging characteristics**
| Variable | STRADL |  | Dutch Famine |  | LBC1936 |  | Simpson's |  |
| --- | --- | --- | --- | --- | --- | --- | --- | --- |
|  | Total<br>N | n (%) | Total<br>N | n (%) | Total<br>N | n (%) | Total<br>N | n (%) |
| Visual ratings |  |  |  |  |  |  |  |  |
| Total SVD score | 1058 |  | 114 |  | 680 |  | 96 |  |
| 0 |  | 461 (43.6) |  | 52 (45.6) |  | 302 (44.4) |  | 12 (12.5) |
| 1 |  | 414 (39.1) |  | 35 (30.7) |  | 249 (36.6) |  | 53 (55.2) |
| 2 |  | 145 (13.7) |  | 16 (14.0) |  | 98 (14.4) |  | 20 (20.8) |
| 3 |  | 31 (2.9) |  | 9 (6.0) |  | 27 (4.0) |  | 8 (8.3) |
| 4 |  | 7 (0.7) |  | 2 (1.3) |  | 4 (0.6) |  | 3 (3.1) |
| Mod/sev total SVD score | 1058 | 188 (17.3) | 114 | 27 (23.7) | 680 | 129 (19.0) | 97 | 31 (32.0) |
| Mod/sev WMH | 1075 | 114 (10.6) | 118 | 30 (25.4) | 685 | 154 (22.5) | 110 | 27 (24.6) |
| Mod/sev EPVS | 1063 | 511 (48.1) | 114 | 28 (24.6) | 680 | 276 (40.6) | 110 | 83 (75.5) |
| 1+ Lacune | 1076 | 86 (7.2) | 118 | 26 (22.0) | 680 | 33 (4.9) | 110 | 27 (24.5) |
| 1+ CMB | 1074 | 119 (11.1) | 117 | 16 (13.7) | 680 | 79 (11.6) | 97 | 11 (11.3) |
| Imaging evidence of 1+ infarcts | 1076 | 60 (5.0) | 118 | 22 (18.6) | 685 | 99 (14.5) | 110 | 10 (7.7) |
| Mod/sev atrophy | 1076 | 70 (5.8) | 118 | 23 (19.5) | 685 | 189 (27.6) | 110 | 64 (58.2) |
| Brain volumes |  |  |  |  |  |  |  |  |
| Whole brain volume (mm3), mean (SD) | 882 | 1,064,225.8 (107,249.4) | - | - | 657 | 990,322.7 (89,401.9) | 110 | 1,137,480.3 (98,056.8) |
| ICV (mm3), mean (SD) | 893 | 1,376,151.5 (226471.3) | - | - | 659 | 1,438,223.1 (133,870.1) | 95 | 1,454,751.50 (123,117.80) |
| WMH volume (mm3), median (IQR) | 471 | 1,510.0 | - | - | 656 | 7,896.0 (11,531.0) | 107 | 25,755.4 (27,166.0) |

Mod/sev WMH= periventricular WMH with a score of 3 and/or deep WMH with a score of 2-3 on the Fazekas scale<sup>34</sup>; mod/sev EPVS= moderate or severe Enlarged Perivascular Spaces; a score of 2-3 on a semi-quantitative scale in the basal ganglia<sup>35</sup>. CMB= cerebral microbleed

Differences in demographic and key characteristics between those who underwent MRI and those who did not are provided in the Supplementary materials and Supplementary Tables 3-6, along with comparisons between the participants in this study and previous waves of each. Where data were available in comparable format, we have also provided key characteristics of the wider Scottish and Dutch population in Supplementary Tables 3-6.

Gender differences were observed in some markers of SVD. Moderate to severe SVD and WMH burden were more common in females compared to males in the LBC1936 (SVD: 22.4% vs 15.9%, *χ*^2^(1)= 4.7, p=0.03; WMH: 26.5% vs 18.8%; (*χ*^2^(1)= 5.8, p=0.02) and Dutch famine birth cohort (SVD: 31.3% vs 14.0%, *χ*^2^ (1)= 4.6, p=0.03). Atrophy was more common in males compared to females in STRADL (11.2% vs 3.1%, *χ*^2^(1)=28.0, p<0.001), the LBC1936 (60.7% vs 40.7%, *χ*^2^(1)= 27.1, p<0.001) and Simpson’s cohort (36.4% vs 18.2%, *χ*^2^(1)= 4.2, p=0.04). No other gender differences were observed in SVD burden.

Results from our main analyses are given below. Analysis of ponderal index are detailed in the Supplementary materials and Supplementary Fig. 2..

### Birth weight

Across all four cohorts, each increase in birth weight of 100g was associated with fewer lacunes (*OR*=0.93 *95%CI*=0.88-0.99), fewer infarcts (*OR*=0.94 *95%CI*=0.89-0.99) and decreased moderate-severe PVS (*OR*=0.95 *95%CI*=0.91-0.99, Fig. 2A) independent of age, sex, hypertension, smoking behaviour and adult SES. Results for the remaining lesions were in the expected direction (increasing birth weight and lower risk of SVD features) but did not reach significance.

**Figure 2.**
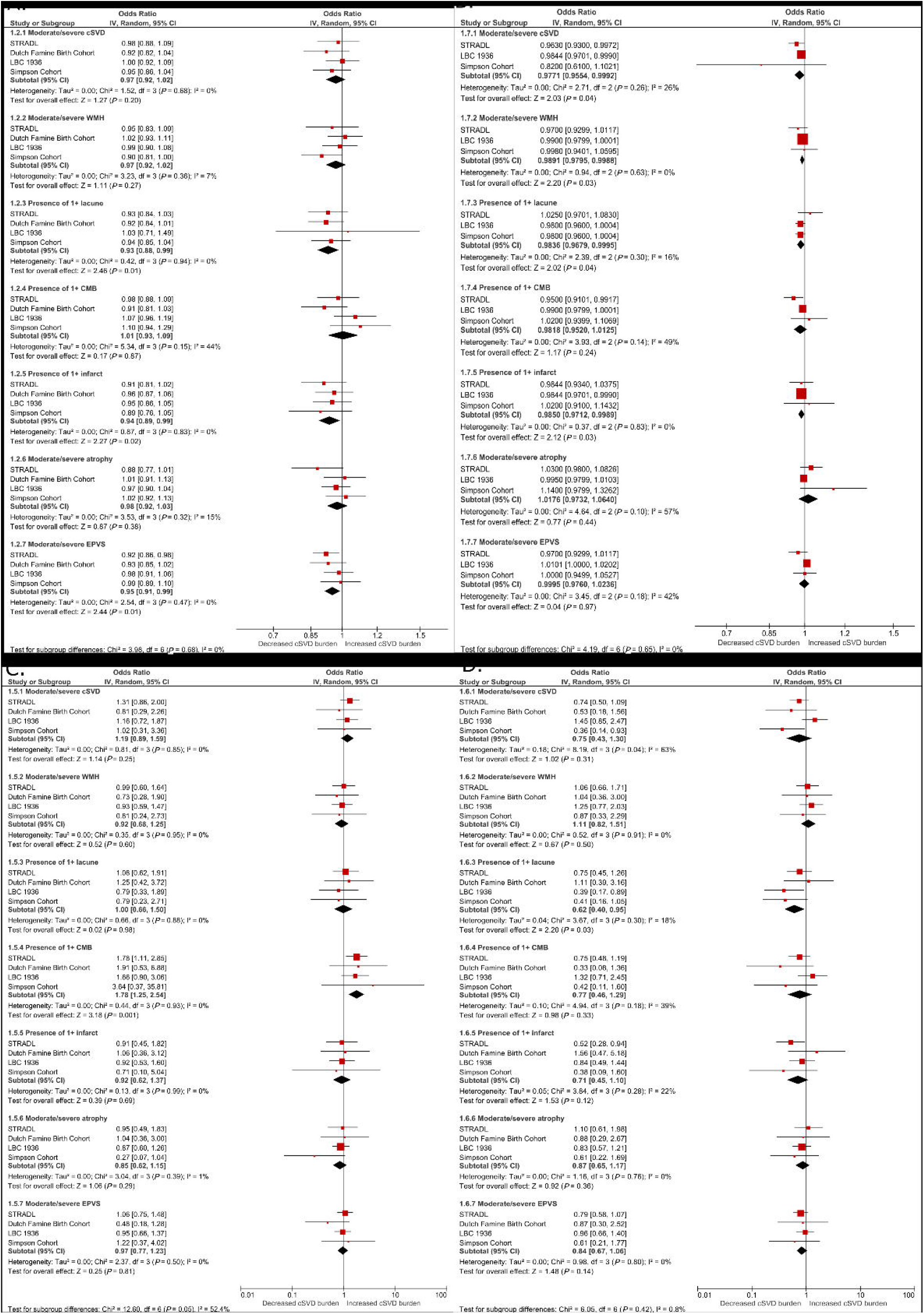
(A-D) Forest plots showing associations between features of SVD and (a) birth weight (b) childhood IQ (c) Low education (d) Low childhood SES. All analyses are adjusted for age, sex, hypertension, smoking behaviour and adult SES.

Associations were attenuated but remained significant after additional adjustment for education and childhood SES (lacunes *OR*=0.94, *95%CI*=0.89-0.99; infarcts *OR*=0.94 *95%CI*=0.89-1.00; PVS *OR*=0.95 *95%CI*=0.91-0.99, Supplementary Table 7).

Increasing birth weight was not associated with WMH volume or brain volume in the Dutch Famine Birth cohort, LBC1936 or Simpson’s cohort (Supplementary Table 8).

### Childhood IQ

Across STRADL, LBC1936, Simpson’s, each point increase in IQ assessed in childhood was associated with decreased risk of moderate or severe WMH (*OR* per point increase 0.99 *95%CI*=0.98-1.00), lacunes (*OR*=0.98 *95%CI*=0.97-0.99), infarcts (*OR*=0.98 *95%CI*=0.97-1.00), and total SVD burden (*OR*=0.98 *95%CI*=0.96-1.00, Fig. 2B) independent of age, sex, hypertension, smoking behaviour and adult SES.

Additional adjustment for education and childhood SES attenuated all associations between childhood IQ and individual SVD features (Supplementary Table 9), but the associations with total SVD burden (*OR*=0.98, *95%CI*=0.97-0.997) and infarcts (*OR*=0.98, *95%CI*=0.97-1.00, Supplementary Table 9) remained.

### Education

Across all cohorts, low education was associated with increased risk of micro-bleeds (vs high education level, *OR*=1.90 *95%CI*=1.33-2.72, Fig. 2C) independent of age, sex, hypertension, smoking behaviour and adult SES. This was attenuated by additional adjustment for childhood IQ and SES (*OR*=1.24, *95%CI*=0.71-2.18, Supplementary Table 9) but remained. The Simpson’s cohort were not included in this multiple regression analysis due to the small number of participants with childhood IQ scores.

Low education was associated with lower brain volume (*MD*= -178.86cm^3^, *95%CI*=-325.07--32.66, Supplementary Fig. 5a) but this was attenuated after adjustment for vascular risk factors and adult SES (β= 0.01, *95%CI*= -0.04-0.06, Supplementary Table 10).

### Childhood SES

Across all cohorts manual childhood SES (i.e. more deprived) was associated with decreased risk of lacunes (*OR*=0.62 *95%CI*=0.40-0.95, Fig. 2D).

## Discussion

Early life factors are thought to influence health later in life but there are few studies with such a wealth of data from birth, childhood and later life to tease out which early life factors are important and if they are independent of each other and of exposures in later life. By combining data from almost 2000 participants from four prospective birth cohorts we confirm that low birth weight, low childhood IQ and less education increase SVD burden 5-8 decades later. SVD is important since it increases dementia and stroke risk, two of the largest sources of loss of independence, health and societal costs in older age across the world. Dementia and stroke prevention are government priorities. Life-course models are increasingly recognised in dementia prevention^36^ but have largely been ignored in stroke and SVD, which too often focus on mid to later life only, thereby missing major opportunities to prevent these devastating diseases much earlier, as well as gaining other health benefits.

Our findings confirm previous findings that some early life factors may increase risk of SVD burden in later life, but importantly also demonstrate that the associations are independent of vascular risk factors and adult SES and persist after adjustment for the other early life factors. Lower birth weight increased the risk of lacunes, infarcts and PVS across four cohorts, independent of education and childhood SES. In STRADL, the LBC1936 and Simpson’s cohort, higher childhood IQ was associated with fewer infarcts and lacunes, lower WMH and total SVD burden. Associations between childhood IQ, infarcts and total SVD burden were independent of education and childhood SES. Across all cohorts, low education level was associated with more micro-bleeds. These new data show that lower birth weight, childhood IQ and low education are independently associated with increased SVD lesions many decades later.

Low childhood SES was not found to be associated with SVD and associations between childhood SES and lacunes were in the opposite direction to what we expected. This was true for univariate analyses (Supplementary Table 8) and multivariate analyses. This may be because childhood SES reflects SES in adulthood, whereas the other early life factors such as cognitive ability and education capture different aspects of early life adversity. Alternatively, parental occupation, which we used as a measure of SES to allow direct comparison between cohorts, may not have been a sufficiently sensitive measure of actual SES in childhood. Jobs traditionally classed as ‘manual’ such as farmer or skipper trawler can have a high income and the wartime occupations of the parents of some cohort members would have been limited. In the LBC1936 we have previously shown a trend towards an association between SVD at age 72 and age 11 deprivation index.^37,38^ Deprivation index encompasses several socioeconomic markers so may be a better measure of SES and thus of associations with SVD in later life.

Increasing age and traditional vascular risk factors, particularly hypertension, are important risk factors for SVD^1,39^ but together explain little variance in WMH (∼2%)^40,41^ suggesting that other factors, as identified here, may contribute to SVD pathology. The effect sizes are small when considered per point difference in IQ score or per 100g difference in birth weight, and the early life variables examined here only explained ∼1% of the variance in SVD risk. However, the fact that these effects are evident for such small differences in scores or weights, and at up to seven decades later, underscores that factors influencing early stages in life, including during foetal development and childhood, can impact on brain health in older age and are rightly public health priorities. Furthermore it is likely that our effects are an underestimate of population effects given that our cohorts are healthier with higher IQ than average members of the population. For example the mean age 11 IQ score of the LBC1936 was relatively high with a narrow range compared with the mean age 11 IQ for Scotland in 1947.^42^

Our associations between birth weight and SVD are independent of gestational age and therefore reflect the impact of variations in growth rather than prematurity. The relationship between size at birth and brain structure is biologically plausible: lack of nutrients at particular stages of gestation can impair foetal growth resulting in small size at birth, indicated by low birth weight or disproportionate growth such as low birth weight to length ratio (ponderal index). Long lasting physiological changes in the structure of foetal organs and tissues can increase risk of later disease in adulthood.^43,44^ Relations between size at birth and disease in later life including coronary heart disease (CHD)^45,46^ are well established, but fewer studies have examined brain health, particularly with this sample size or age range. The current study is one of the few examining the effect of size at birth on brain volumes in later life and the first to examine multiple markers of SVD.

We found no associations between birth weight or ponderal index and WMH burden or brain volumes. This is consistent with data from the (AGES)-Reykjavik study (RS)^47^ which reported no association between ponderal index and WMH burden at age 75 after adjustment for vascular risk factors. Birth weight and size are indirect measures of the fetal environment and may not reflect all adverse prenatal circumstances that can affect later life health. The Dutch Famine Birth Cohort previously showed that foetal malnutrition can lead to accelerated cognitive ageing and advanced structural brain ageing, measured using the BrainAGE method (a composite measure based mainly on tissue loss) independent of birth weight.^48^

From a life course perspective, a disadvantaged foetal environment may interact with factors during childhood to increase risk of later disease. Development of neural pathways in the brain extends well into childhood and may therefore mean the brain remains vulnerable to insults for a longer period of time.^49^ Our two recent meta-analyses^14,50^ found small but statistically significant associations between increasing childhood IQ and lower WMH burden (*r* = -0.07) and a 17% lower risk of stroke. Low education (defined by attainment or years) was associated with a 35% relative increased risk of stroke and a 17% increased risk of SVD. Manual paternal occupation (SES measure) was associated with a 28% increased risk of stroke and increased WMH (only one study identified). However, the previous literature did not allow us to determine the independent effect of these three inter-related early life factors from each other, or from risk factor exposures in adulthood, which we are now able to do.

In many high-income countries age specific incidence rates of dementia are declining.^51,36^ Improved health in old age, including cerebrovascular disease and SVD^52^, has been reported across generations and epidemiological studies have found that age adjusted incidence rates of dementia are lower in more recent cohorts compared with cohorts from previous decades.^36,51,52^ This can in part be attributed to population public health strategies, advances in treatment and management of patients with cerebrovascular disease and dementia, and improved management of key modifiable risk factors such as smoking and hypertension. Additionally, investment in early life, particularly improvements in living conditions and education, explain some of the decline in incidence of dementia.^53-55^ More recent generations of older adults have received more years of statutory education than older cohorts which may increase cognitive reserve and therefore reduce risk of dementia or cerebrovascular disease. This is particularly relevant to our cohorts, whose years of birth span the 20^th^ century. Low education increased with increasing age of our cohorts, as did SVD burden. In STRADL (median year of birth 1955) 24% had low education and 17.3% had moderate to severe SVD burden. In the Simpson’s cohort (born 1921-1926) 81% had low education and 32% had moderate to severe SVD burden. Increases in life expectancy means that the global population is aging, therefore identifying factors that contribute to reductions in the prevalence and incidence of dementia and cerebrovascular disease is a major priority. Our findings support the suggestion that reducing inequalities, including improvements in education, will contribute to improvements in health in older age and a reduction in the risk of dementia and cerebrovascular disease.

Why might the early life factors increase the risk of SVD in later life? There are numerous potential explanations. Children with higher IQ or from higher socioeconomic backgrounds are likely to receive better diets, medical care, more educational opportunities and hence better job opportunities or less hazardous working conditions. In adulthood, they may be more likely to engage in better lifestyle behaviours and self-management of vascular risk factors. Alternatively, positive early life factors may be associated with, or lead to, an increase in the resilience and integrity of the brain resulting in less SVD. These remain important empirical questions to be addressed in future work.

## Strengths and limitations

Strengths include data collected prospectively in early life through to middle or later life, including brain imaging, from different studies in two western European countries. Detailed birth records allowed correction for gestational age and did not rely on retrospective estimations of birth weight. We used ponderal index and birth weight as measures of infant growth. Ponderal index may be a better indicator of gestational problems than birth-weight percentiles as it provides information on the neonate’s body proportionality and can detect situations in which weight growth exceeds or fails to match growth in the infant’s length.^56^ We adjusted for key adult vascular risk factors and other early life factors in our analyses with a relatively large sample size for some analyses. We also did a detailed characterisation of SVD using multiple individual assessments as well as a summary score.

Limitations include availability of birth data only for some participants in STRADL and the LBC1936. Participants in the Dutch Famine Birth Cohort may be unusual due to their famine exposure, and we have demonstrated excess mortality up to the age of 63 years in women exposed to famine in early gestation^57^. This may have resulted in selective participation of people who were in sufficient health to participate in the present study at age 68 years. Participants with birth data were born in hospitals, which was uncommon at the time of their births. In the Netherlands women largely delivered at home supported by a midwife. Whilst little is known about the actual referring pattern during this period most referrals to hospital were because of social or medical reasons and most referred women were from lower or middle social classes. Two of our cohort’s early childhood or early adulthood were spent during the Second World War, which may have influenced the development of cognitive ability or educational opportunities. Although this seems unlikely as IQ scores of those who took the Moray House Test No. 12 in 1947 (born 1936) were higher than the cohort who took them in 1932 (born 1921). The four cohorts recruited community-dwelling volunteers who may be healthier, with less socioeconomic adversity than non-volunteers. Within our cohorts those who completed the MRI were younger and healthier than those who declined. Participants in all but one cohort were largely female and when compared to the Scottish and Dutch population had lower risk factor profiles, were more educated and from higher adult socioeconomic class. Even in our oldest cohort aged 80 years, less than 30% of participants had moderate or severe SVD. The large sample size of some of our cohorts mean that there are participants with a range of socioeconomic backgrounds and medical conditions, but our samples may not be truly representative of the populations from which they are drawn. Our samples came from three regions of Scotland and one region of the Netherlands, which may introduce effects due to local variations in socioeconomic strata but may also increase the generalisability of our findings and may also be considered a strength of our study. Years of education were not available for all cohorts and the education system in the Netherlands differs from that in Scotland which meant the division into ‘low’ and ‘high’ education level was relatively crude. Whilst we adjusted our models for key vascular risk factors, it was not possible to separate the confounding effects of other prenatal environmental or genetic influences which may affect foetal brain development. In this study we did not adjust for multiple comparisons as a Bonferroni-style correction would have been inappropriate when our variables are not independent. We dealt with multiple comparisons as recommended by Perneger^58^ by transparently reporting all results, including those with borderline significance. We also specified our hypotheses a priori based on previous research. However, given the number of statistical comparisons in our analysis it is still possible that some of our associations may be due to Type I error.

SVD frequently coesixtis with neurodegenerative disease. We did not examine associations between early life factors and biomarkers such as amyloid-β, tau or synclein but given the overlap between neurodegenerative and cerebrovascular pathologies, including shared risk factors^59^, it is possible that the associations observed here may interact with degenerative neuropathologies.

## Conclusions

Our findings suggest an important effect of early life factors, particularly childhood IQ, on brain vascular disease in later life, independent of common vascular risk factors, adult SES and other early life factors. Positive early life factors may influence health behaviours and access to socioeconomic resources beneficial to health, or may increase brain integrity and resilience reducing susceptibility to cerebrovascular disease. Brain vascular disease increases the risk of cognitive impairment, dementia and stroke^1^ and worsens chances of recovery after stroke.^3^ The current findings may provide a possible mechanistic link between early life factors and risk of stroke and dementia. Health disparities are well known and these findings suggest that such disparities may have effects persist across more than seven decades of life, highlighting the importance of identifying modifiable early life factors as targets for future social policy interventions with have long-lasting impacts.

## Data Availability

The data that support the findings of this study are available upon reasonable request.

## Abbreviations

ACDS: Aberdeen Child Development Survey
ACONF: Aberdeen Children of the 1950s cohort
DOHAD: Developmental Origins of Adult Heath and Disease
GS:SFHS: Generation Scotland: Scottish Family Health Study
ICV: intracranial volume
LBC1936: the Lothian Birth Cohort 1936
LGA: Lesion Growth Algorithm
PVS: perivascular spaces
SES: socioeconomic status
STRADL: STratifying Resilience and Depression Longitudinally
SVD: cerebral small vessel disease
WMH: white matter hyperintensities

