## supplementary for "Early life predictors of late life cerebral small vessel disease in four prospective cohort studies"

### Supplementary materials

#### Supplementary figure 1: Recruitment flow chart of STRADL (A), the Dutch Famine Birth Cohort (B), LBC 1936 (C) and the Simpson's cohort (D).

##### A. STRADL

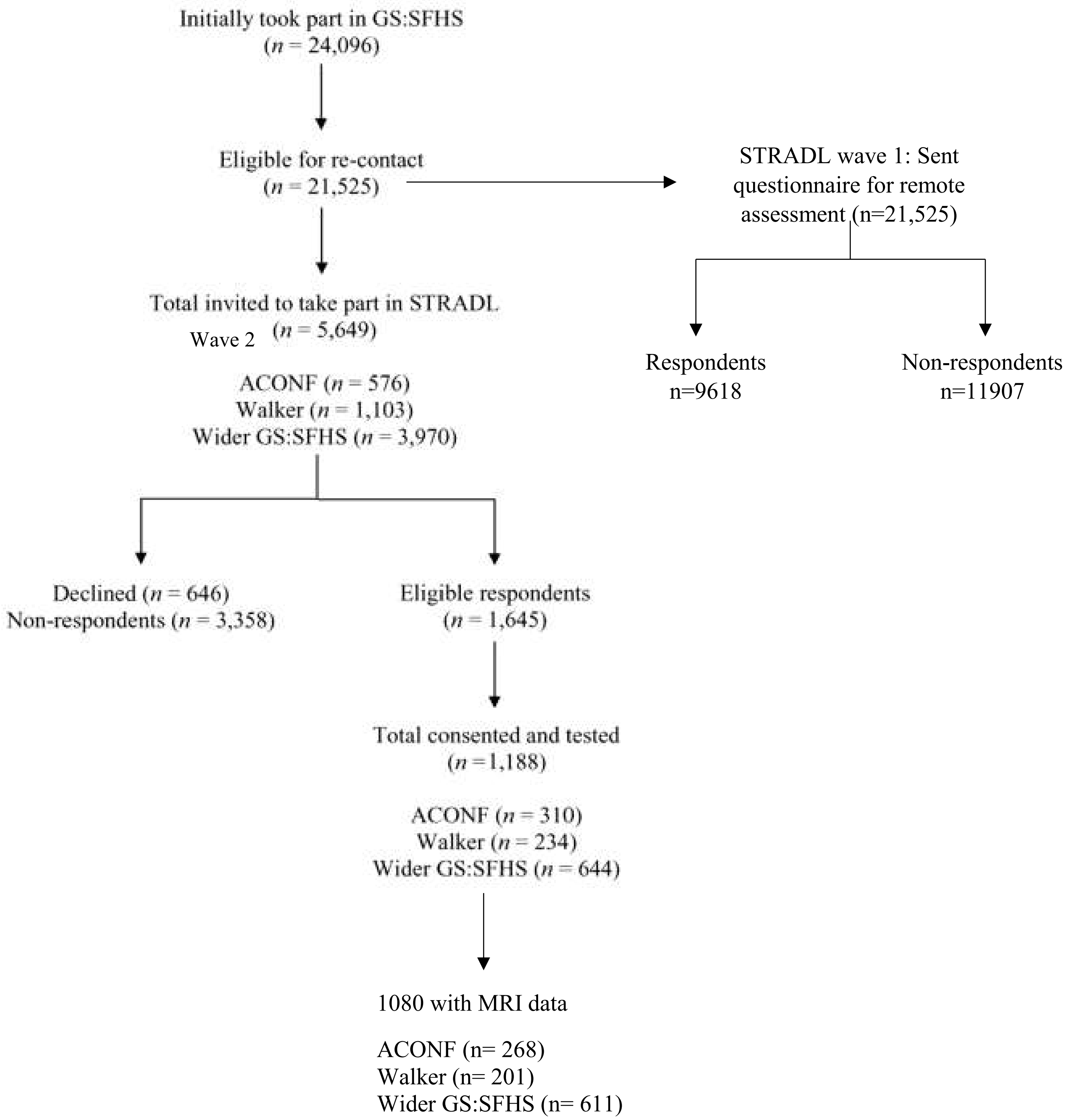


GS:SFHS= Generation Scotland: Scottish Fam [illegible]

ACONF= Aberdeen Children of the 1950's cohort.

Adapted from: Habota T, Sandu AL, Waiter GD et al. Cohort profile for the STratifying Resilience and Depression Longitudinally (STRADL) study: A depression-focused investigation of Generation Scotland, using detailed clinical, cognitive, and neuroimaging assessments [version 1]. Wellcome Open Res 2019, 4:185 (doi: 10·12688/wellcomeopenres.15538·1)

#### B. The Dutch Famine Birth Cohort

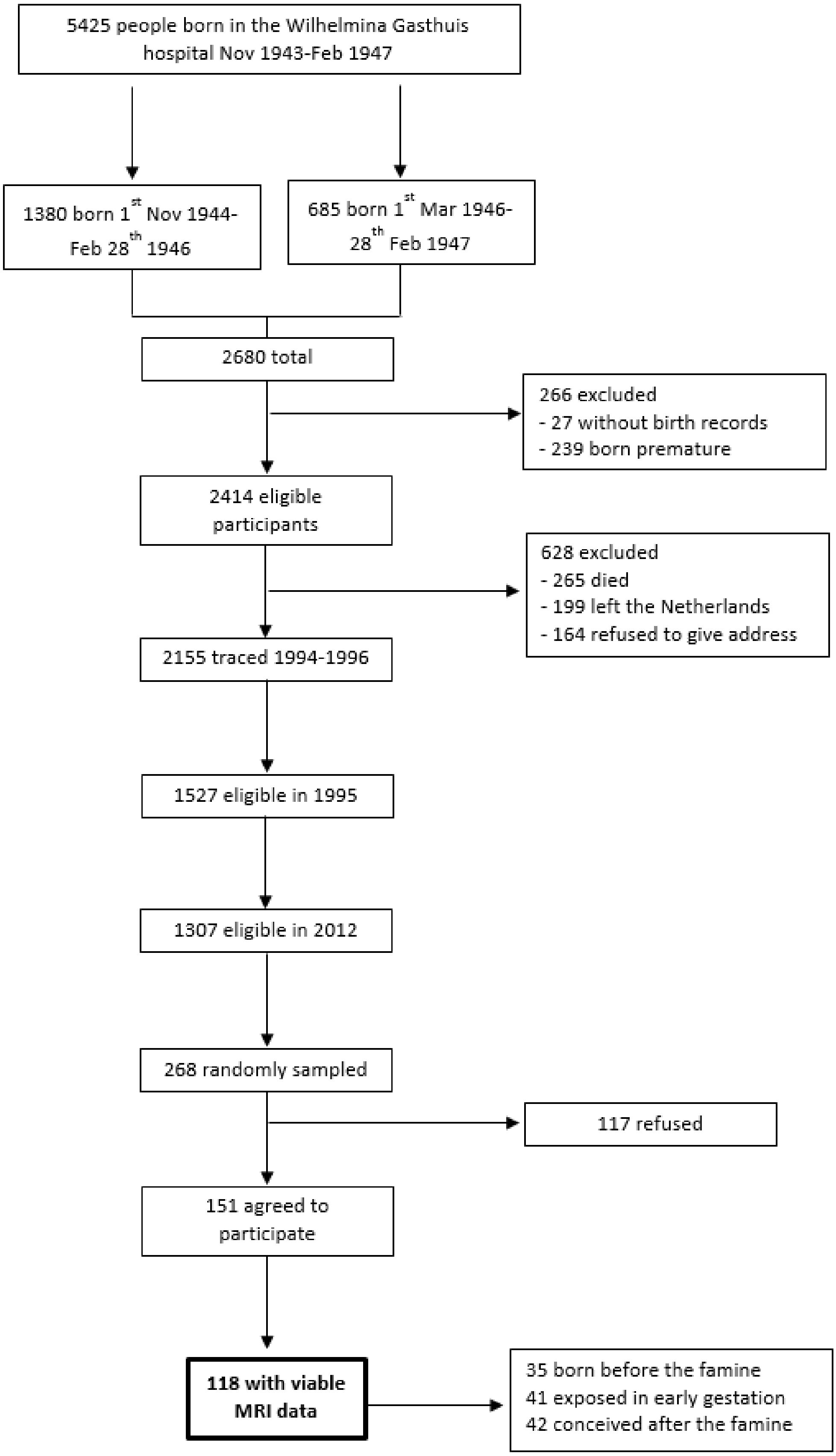

#### C. The Lothian Birth Cohort 1936

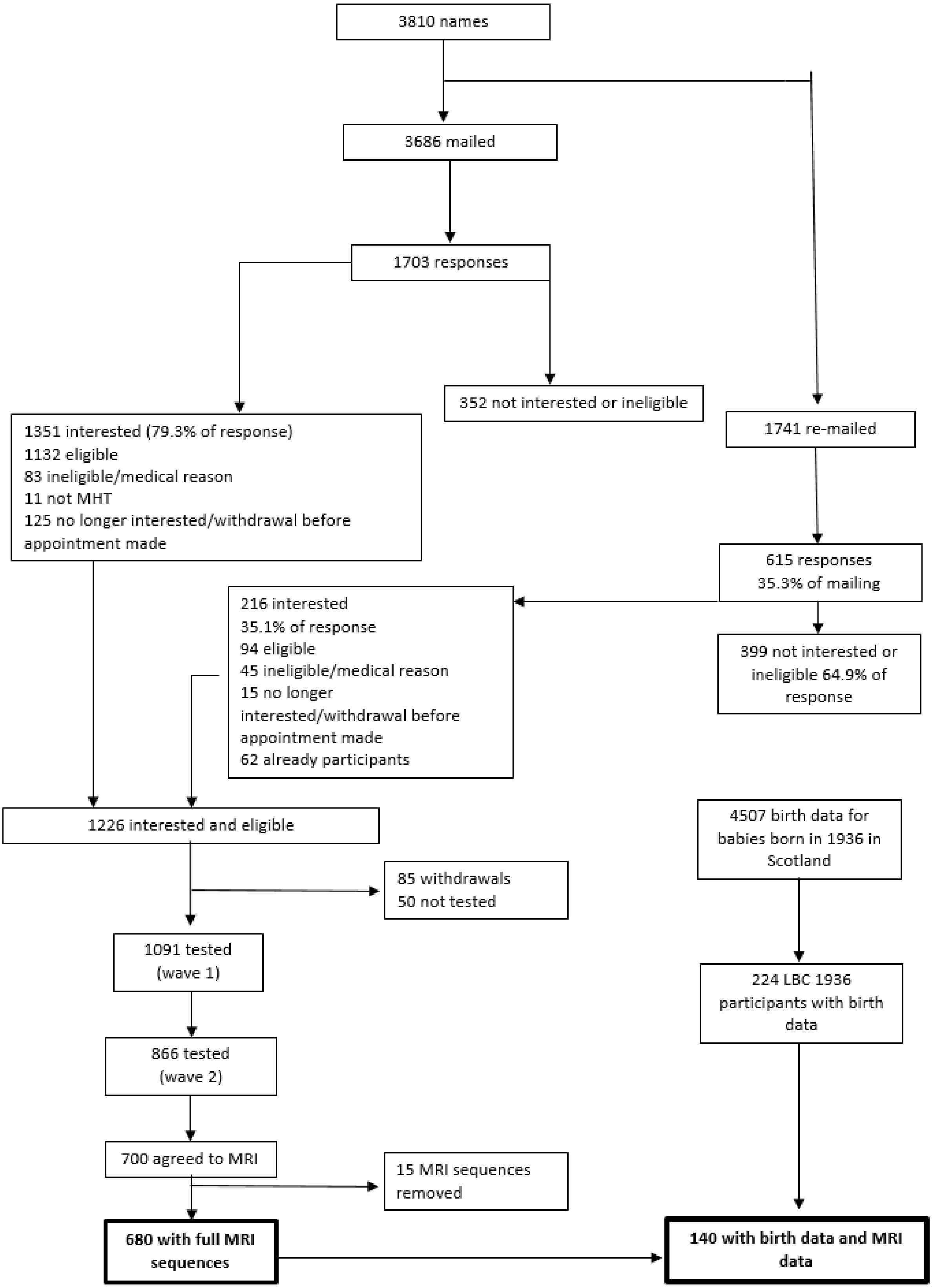


Adapted from: Deary, BMC Geriatrics 2007, 7:28

#### D. The Simpson's Birth Cohort Study

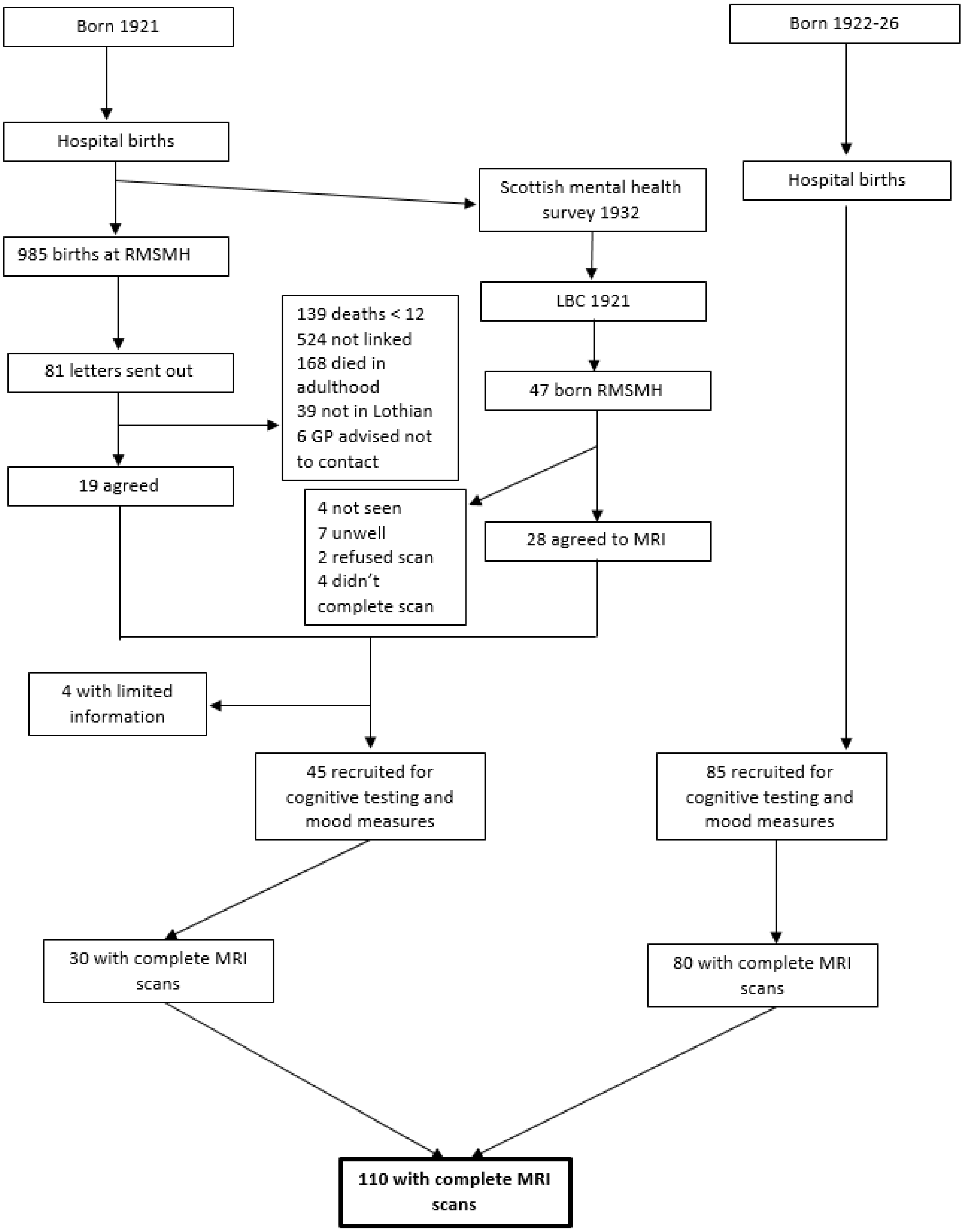


RMSM= Royal Maternity and Simpson Memorial hospital

Adapted from: Shenkin, SD (2006) *Life course influences on cognitive ability and cerebrovascular disease* (MD thesis)

#### Supplementary table 1: Details of early life factors available in all cohorts

| Early life factor | STRADL | Dutch Famine Birth Cohort | LBC 1936 | Simpson's cohort | Descriptive statistics |
|---|---|---|---|---|---|
| Birth weight | **Available for:** 1) all ACONF participants 2) 154 Walker participants.<br>**Taken from**: 1). the ACONF database which had birth records taken from the Aberdeen Maternity and Neonatal database at the Aberdeen Maternity Hospital. Recorded in pounds and ounces and converted to grams.<br>2·) the Walker database which had birth records in Dundee. Recorded in grams. | **Available for:** all participants<br>**Taken from**: birth records at the Wilhelmina Gasthuis Hospital in Amsterdam. Recorded in grams | **Available for:** 140 participants<br>**Taken from**: birth records from the Simpson Memorial Hospital, Bellshill Hospital Lanarkshire and Aberdeen Maternity Hospital. Recorded in pounds and ounces and converted to grams. | **Available for:** all participants<br>**Taken from**: birth records from the Royal Maternity and Simpson Hospital, the Elsie Memorial Hospital and the Lyin-in Institution. Recorded in pounds and ounces and converted to grams | Mean (SD)<br>**STRADL:** 2·4% participants with birth weight <5lbs; 3309·3 (SD 529·4)<br>**Dutch Famine Birth Cohort**: 3417·5g (SD 503·4)<br>**LBC 1936:** 3351·5g (SD 482·1)<br>**Simpson's cohort:** 3333·6g (SD 457·2);<br><br>**ANOVA:** F(3, 365)= 0·99, p=0·37 |
| Ponderal index | Not collected | **Available for:** all participants<br>**Calculated from**: birth weight and length as birth weight/birth length$^3$ | **Available for:** 79 participants<br>**Calculated from**: birth weight and length birth weight/birth length$^3$ | **Available for:** all participants<br>**Calculated from**: birth weight and length birth weight/birth length$^3$ | Mean (SD)<br>**Dutch Famine cohort:** 26·16 (SD 2·30)<br>**LBC 1936:** 27·33 (SD 5·29)<br>**Simpson's cohort:** 25·55 (SD 4·10)<br><br>**ANOVA:** F(2, 297)= 4·72, p=0·01 |
| Childhood IQ | **Available for:** all ACONF participants<br>**Calculated by:** adjusting the raw scores from the Schonnell and Adams Essential Intelligence Test (age 9 IQ), for age in days at testing and placed on an IQ type scale (mean=100, SD=15). | Not available | **Available for**: all participants<br>**Calculated by:** adjusting the raw scores from the Moray House Test number 12 (MHT) (age 11 IQ), for age in days at testing and placed on an IQ type scale (mean=100, SD=15). | **Available for:** 47 participants<br>**Calculated by:** adjusting the raw scores from the Moray House Test number 12 (MHT) (age 11 IQ), for age in days at testing and placed on an IQ type scale (mean=100, SD=15). | Mean (SD)<br>**STRADL:** 102·0 (8·9)<br>**LBC 1936:** 100·80 (SD 15·3)<br>**Simpson's cohort:** 101·7 (14·5)<br><br>**ANOVA:** (F(2)= 1·35, p= 0·26 |
| Education | **Available for:** all participants<br>**Measured:** divided into compulsary, more than compulaary and post secondary and dichotomised at compulsary. | **Available for:** all participants<br>**Measured:** a 10 point scale (1= Primary education not completed, 10=University completed) and dichotomised at lower secondary school | **Available for:** all participants<br>**Measured:** using years of education and dichotomised at 11 years. | **Available for:** all participants<br>**Measured:** using years of education and dichotomised at 11 years. | % of participants with low education (≤ 11 years)<br><br>**STRADL** 24%; **Dutch Famine** 62·7%<br>**LBC 1936** 71·7%<br>**Simpson's cohort** 80·9%<br><br>**Chi square:**<br>$\chi^2(3)$= 8·0, p=0·05 |
| Childhood SES | **Available for:** all participants<br>**Obtained from:** self-reports by each participant at the time of the interview in adulthood and classified as manual and non-manual | **Available for:** all participants<br>**Obtained from**: birth records We classified childhood SES as manual and non-manual at the time of the child's birth | **Available for:** all participants<br>**Obtained from:** self-reports by each participant at the time of the interview in adulthood and checked using the General Register Office's Census, 1951. Classification of Occupations We classified childhood | **Available for:** all participants<br>**Obtained from:** self-reports by each participant at the time of the interview in adulthood and classified as manual and non-manual at age 11 | % of participants with low childhood SES (manual parental occupation)<br><br>**STRADL:** 67·2%<br>**Dutch famine birth cohort:** 66·7%<br>**LBC 1936:** 74·2%<br>**Simpson's cohort:** 69·1% |

| | | | | | |
|---|---|---|---|---|---|
| | | | SES as manual and non-manual at age 11 | | **Chi square:** $\chi^2(3)= 5{\cdot}7$, p=0·13 |

#### Supplementary Table 2: Sequence parameters for MRI scanning in all cohorts

| Sequence name | Acquisition method | Field of view (mm) | Matrix | Slices | Thickness (mm) | Voxel (mm) | TR/TE/TI (ms) |
|---|---|---|---|---|---|---|---|
| **STRADL (Aberdeen)** | | | | | | | |
| T2-weighted | 3D TSE | 250 | 512 x 512 | 320 | 0·5 | 0·5 x 0·5 x 0·5 | 2500/314 |
| SWI | FFE | 230 | 768 x 768 | 130 | 1 | 0·3 x 0·3 x 1.0 | 31/7·2,14·4,20·6,26·8 |
| FLAIR | 3D IR FSE | 240 | 256 x 256 | 160 | 1 | 0·94 x 0·94 x 1.0 | 8000/349/2400 |
| T1-weighted volume | 3D IR TFE | 240 | 256 | 160 | 1 | 1.0 x 1.0 x 1.0 | 8·3/3·8/1031 |
| **STRADL (Dundee)** | | | | | | | |
| T2-weighted | 3D TSE | 256 | 256 x 256 | 320 | 0·5 | 0·5 x 0·5 x 0·5 | 3200/ 408 |
| SWI | FFE | 230 | 320 X 160 | 144 | 1.0 | 0·4 x 0·4 x 1.0 | 28/20 |
| FLAIR | 3D IR FSE | 256 | 256 X 256 | 160 | 1.0 | 1.0 x 1.0 x 1.0 | 5000/386/1800 |
| T1-weighted volume | 3D IR TFE | 256 | 256 x256 | 208 | 1.0 | 1.0 x 1.0 x 1.0 | 1740/ 2·62/ 900 |
| **Dutch Famine Birth cohort** | | | | | | | |
| T2-weighted | 2D TSE | 230 x 230 | 512 x 512 | 28 | 4 | 0·45 x 0·45 | 3000/80 |
| SWI-weighted | 3D SWI | 220 x 220 | 448 x 448 | 220 | 1.2 | 0·49 x 0·49 x 0·6 | 19·8/25·7 |
| FLAIR | 3D | 250 x 250 | 240 x 240 | 321 | 1.1 | 1.1 x 1.1 x 1.12 | 4800/365 |
| T1-weighted volume | 3D TFE | 256 x 256 | 256 x 256 | 180 | 1.0 | 1.1 x 1.1 x 1.2 | 6·8/3·1 |
| **LBC 1936** | | | | | | | |
| T2-weighted | FSE | 256 x 256 | 256 x 256 | 80 | 2 | 1 x 1 x 2 | 1 1320/105 |
| T2*-weighted | Gradient echo | 256 x 256 | 256 x 192* | 80 | 2 | 1 x 1 x 2 | 940/15 |
| FLAIR | FSE | 256 x 256 | 256 x 192* | 40 | 4 | 1 x 1 x 4 | 9002/147/2200 |
| T-1 mapping | FSPGR | 256 x 256 | 128 x 128* | 72 | 2 | 1 x 1 x 2 | 6/2 |
| T1-weighted volume | 3D IR-Prep FSPGR | 256 x 256 | 192 x 192* | 160 | 1.3 | 1 x 1 x 1.3 | 10/4/500 |
| **Simpson's cohort** | | | | | | | |
| T2-weighted | FSE | 240 x 240 | 256 x 256 | - | 5 | - | 6300/102 |

| | | | | | | | |
|---|---|---|---|---|---|---|---|
| T2*-weighted | Gradient echo | - | - | - | - | - | - |
| FLAIR | FSE | 240 x 240 | 256 x 192 | - | 5 | - | 9000/140/2200 |
| T-1 mapping | FSPGR | 240 x 240 | 256 x 224 | - | 5 | - | 450/8 |
| T1-weighted volume | 3D IR-Prep FSPGR | 240 x 240 | 256 x 256 | - | 1.7 | - | 400 |

* Zero filled to 256 x 256; TR: repetition time; TE: echo time; FLAIR: Fluid-attenuated inversion recovery; SWI: susceptibility weighted imaging; IR-Prep: inversion recovery prepared; FSPGR: fast spoiled gradient echo; FSE: fast spin echo. – is used where data were not available

Comparison of participants with and without MRI.

Those who declined MRI were older (STRADL U= 54087.5, p=0.01; Dutch Famine Birth cohort (U= 1353.5, p=001); Simpson's cohort (U=530.5, p<0.001) and more likely to be diagnosed hypertensive (Simpson's cohort $\chi^2$ (1)= 4.4, p=0.036) and diabetic (STRADL only $\chi^2$(1)= 14.4, p<0.001). In STRADL and the LBC1936 they were also more likely to have lower childhood IQ (STRADL U= 1947.5, p=0.03; LBC1936 U= 110284.0, p=0.01) and lower adult and childhood social class (STRADL only; manual vs non-manual adult occupation: $\chi^2$ (1)=9.2, p<0.01), manual vs non-manual Father's occupation $\chi^2$ (1)= 4.5, p=0.04).

Comparison of participants and non-participants.

Participants in this wave of the Dutch Famine Birth cohort were randomly sampled from 1307 eligible cohort members. Participants in this study had a higher birth weight than eligible non-participants (3438g vs 3346g). In STRADL those who participated in the face to face assessment in this study were older (median age 62) with higher levels of education (40% with university education) compared to the previous wave of STRADL and the GS:SFHS population. In the previous wave of STRADL respondents to the remote assessment were predominantly female (62%) and were older (mean 50.48 SD 13.41) and from less socioeconomically deprived areas compared with non-respondents in the Scottish Index of Multiple Deprivation (SIMD) 2009 (4123 vs 3733)[15]. Participants in this this wave of the LBC1936 were less likely to be hypertensive, have high cholesterol and smoke at wave 1 than those who declined to participate and less likely to have low education and childhood IQ.

**Supplementary Table 3: Key characteristics of STRADL face-to-face assessment broken down by MRI status, STRADL remote follow up, wider GS:SFHS baseline assessment and the wider Scottish population.**

| | | | STRADL | | | | |
|---|---|---|---|---|---|---|---|
| | Scottish Population[60,61] | GS:SFHS total (N=21525)[62] | Wave 1 Remote assessment | | **This study Face-to face assessment (n= 1,188)** | | |
| | | | Non-respondents (N=11907) | Respondents (N= 9618) | All (n= 1,188) | With MRI (n=1076) | Without MRI (n=109) |
| Gender (n, % female) | 51.5 | 41 | 57 | 62§ | 59 | 40.5 | 50.5 |
| Mean age (SD) | - | - | - | - | 59.3 (10.1) | 59.3 (10.2) | 62.1(8.7)* |
| Median age Male | - | 47 | 43 | 54 | 62 | 62 | 62 |
| Median age Female | - | 48 | 45 | 52 | 61 | 61 | 61 |
| Hypertension, % | 66 | 27.2 | - | - | 27.7 | 27.7 | 33.6 |
| Diabetes, % | 5 | 6.9 | - | - | 7.9 | 6.4 | 16.0** |
| Current smokers % | 21 | 35 | - | - | 40.3 | 45.3 | 44.6 |
| Annual income >£30,000 (%) | | 60 | 57 | 63 | - | - | - |
| SIMD | | 3910 (1842) | 3733 (1875) | 4123 (1777) | - | - | - |
| Adult SES, manual | - | - | - | - | 31.9 | 32.2 | 46.4** |
| Employment in those aged up to 75 years (%) | | | | | | | |
| Unemployed | 5 | 1.7 | 5 | 4 | 3 | - | - |
| Retired | 12.9 | 15.1 | 13 | 18 | 32 | - | - |
| Employed | 69 | 62.8 | 71 | 71 | 65 | - | - |
| Education (%) | | | | | | | |
| Degree | 26 | 33 | 28 | 37 | 40 | - | - |
| No qualifications | 27 | 5 | 9 | 7 | 25 | - | - |
| Childhood IQ, mean (SD) | - | - | - | - | 102.0 (8.9) | 102.5 (8.8) | 97.4 (12)* |
| Low education, n (%) | - | - | - | - | 24.0 | 24.0 | 29.7 |

| | | | | | | | |
|---|---|---|---|---|---|---|---|
| Manual childhood SES, % | - | - | - | - | 67.2 | 67.2 | 77.1* |
| Birth weight, g, mean (SD) | - | - | - | - | 3309.3 (529.4) | 3309.3 (529.4) | 3309.3 (436.5) |

§ t(21457)= -31.25, p<0.001, cohen's d=0.21)[15]; * those with MRI vs those without MRI p<0.05; ** those with MRI vs those without MRI p<0.01
Abbreviations: SIMD the Scottish Index of Multiple Deprivation 2009: numbers represent mean rank (SD); GS:SFHS Generation Scotland Scottish Family Health Study; SES socioeconomic status; - Is used where data are not available.

**Supplementary Table 4: Key characteristics of the Dutch Famine Birth cohort in the current study broken down by MRI status, previous waves and the wider Dutch population.**

| | | | 1994-1996 (Aged 50)[67] | 2002-2004 (Aged 58)[68] | This study 2012-2013 (Aged 68) | | |
|---|---|---|---|---|---|---|---|
| | Dutch population[63,64,65,66] | Original cohort (n=2414)[67] | (n=741) | (n=721) | All (n=151) | With MRI (n=118) | Without MRI (n=33) |
| Age, mean (SD) | | | | | 67.5 (0.9) | 67.5** (0.9) | 68.0** (0.8) |
| Females, % | 50.5 | 48.6 | 52 | 53 | 55.6 | 55.9 | 54.5 |
| Adult SES, manual, % | - | | | | 41.1 | 37.3 | 54.5 |
| Birth weight (g), mean (SD) | - | 3346 (478) | 3347 | 3363 (465) | 3438.3* (488.0) | 3417.5 (503.4) | 3512.4 (427.5) |
| Ponderal index, mean (SD) | - | 26.4 (2.4) | 26.3 (2.3) | 26.3 (2.3) | 26.2 (2.3) | 26.2 (2.3) | 26.7 (2.6) |
| Hypertension, % | 20.1 | - | | | 54.7 | 62 (53) | 20 (60.6) |
| Aged 50-59 | 33.7 | - | | | | | |
| Systolic BP, mean (SD) | | - | 126 (16) | 137 (18) | 151 (18) | 152 (17) | 148 (23) |
| Diabetes, % | | - | | | 19.2) | 20.3 | 15.2 |
| Hypercholesterolimia, % | | - | | | 47.3 | 47.9 | 45.0 |
| Smoking, current, % | 19.0 | - | 34.0 | 23.0 | 11.3 | 11.0 | 12.1 |
| SES (ISEI) | 46 | - | 48 (13) | 47 (40-59) | 51 (16-77) | 51 (16-77) | 43 (25-76) |
| Low education, % | - | - | 66 | | 60.3 | 62 | 52 |
| Low childhood SES, n (%) | | 76.0 | 70.0 | 74.0 | 67.5 | 66.7 | 70.4 |

* Participants in this study vs eligible non-participants p<0.05; Abbreviations: SES socioeconomic status; ISEI A standard international socio-economic index of occupational status (higher number represents higher socioeconomic status).

**Supplementary Table 5: Key characteristics of wave 1, wave 2 and those who refused wave 2 of the LBC1936 compared to the wider Scottish population. Characteristics of participants in wave 2 are further broken down by MRI status.**

| | | Wave 1 | | This study (Wave 2) | | |
|---|---|---|---|---|---|---|
| | Scottish population[61,69] | Wave 1 all (n=1091) | Refused wave 2 (n=225) | All (n=866) | With MRI (n=685) | Without MRI (n=186) |
| Median age | - | 69.5 (0.8) | 69.6 (0.8) | 72.7 (0.7) | 72.7 (0.7) | 72.8 (0.7) |
| Gender (n, % female) | 51.5 | 49.8 | 55.6 | 48.3 | 52.8 | 47.8 |
| Adult SES¥ | - | 2.4 (0.91) | 2.6 (0.86) | 2.4 (0.9) | 2.4 (0.9) | 2.4 (0.9) |
| Adult SES, manual, n (%) | - | 21.7 | 25.5 | 20.7 | 79.6 | 20.4 |
| Current smokers, n (%) | 31.0 | 11.5 | 21.8** | 8.4 | 8.2 | 9.1 |
| Hypertension, n (%) | 41.0 | 433 (39.7) | 103 (45.8)* | 49.1 | 49.1 | 48.9 |
| Diabetes, n (%) | 3.8.0 | 91 (8.3) | 16 (7.1) | 11.0 | 10.3 | 13.4 |
| Hypercholesterolemia, n (%) | - | 386 (35.4) | 93 (41.3)* | 33.9 | 41.9 | 38.2 |
| Childhood IQ, mean (SD) | - | 100.0 (14.9) | 97.4 (13.4)** | 100.7 (15.3) | 100.8 (15.3) | 100.1 (0.6)∞ |
| Education (%) | | | | | | |
| Degree | 20.0 | 158 (14.5) | 25 (11.1) | 15.4 | 16.2 | 12.4 |
| No qualifications | 33.0 | 193 (17.7) | 40 (17.8) | 17.7 | 17.1 | 19.9 |
| Low education n (%) | - | 812 (74.4) | 184 (81.8)** | 72.5 | 71.6 | 75.8 |
| Father's SES¥ | - | 2.91 (0.94) | 2.87 (0.94) | 2.92 (0.9) | 2.9 (0.9) | 2.9 (1.1) |
| Manual childhood SES, n (%) | - | 700 (72.9) | 121 (53.8) | 73.5 | 74 | 71.7 |
| Birth weight (g), mean (SD) | - | 3299.3 (490.5) | 3223.1 (476.8) | 3323.2 (493.7) | 3348.9 (482.9) | 3215.1 (531) |
| Ponderal index, mean (SD) | - | 26.9 (5.0) | 25.8 (4.1) | 27.3 (5.2) | 27.4 (5.3) | 27.2 (4.8) |

* Those who refused wave 2 vs participants of wave 2 $p<0.05$; **Those who refused wave 2 vs participants of wave 2 $p<0.01$; ∞those with MRI vs those without MRI $p<0.05$; Abbreviations: SES socioeconomic status ; ¥ Father's SES and adult SES are based on five categories from professional (1) to unskilled labour (5) (higher number indicates lower SES; - is used when data were not available.

**Supplementary Table 6: Key characteristics of the Simpson's cohort broken down by MRI status, characteristics of the LBC1921 and the wider Scottish population.**

| | Scottish Population[61,69] | LBC1921[17] | Simpson's | | |
|---|---|---|---|---|---|
| | | Wave 1 | All (n=130) | With MRI (n=110) | Without MRI (n=20) |
| Mean age | - | - | 78.4 (30.8) | 78.2(1.4) | 79.5** (1.1) |
| Gender (% female) | 51.5 | 54.0 | 69.2 | 70.0 | 65.0 |
| Adult SES*, mean (SD) | - | 2.24 (0.88) | 2.6 (0.7) | 2.5 (0.8) | 2.9 (0.5) |
| Adult SES, manual, % | - | - | 62.3 | 58.2 | 85.0 |
| Current smokers (%) | 31 | - | 9.2 | 7.3 | 20.0 |
| Hypertension (%) | 41 | - | 48.5 | 44.5 | 70.0** |
| Diabetes (%) | 3.8 | - | 5.4 | 6.4 | 0.0 |
| Childhood IQ | - | 101.6 (14.2) | 100.1 (14.8) | 101.7 (14.50) | 15.4 |
| MHT raw score, mean (SD) | 34.5 (15.5) | - | 42.9 (14.0) | 44.5 (13.7) | 38.8 (14.6) |
| Low education (%) | - | - | 82.3 | 80.9 | 90 |
| Father's SES*, mean (SD) | - | 2.73 (0.93) | 3.2 (0.8) | 3.2 (0.9) | 3.25 (0.8) |
| Father's SES, manual (%) | - | - | 70.0 | 69.1 | 75.0 |
| Birth weight, (g) mean (SD) | - | - | 3331.5 (461.7) | 3333.6 (457.2) | 3319.65 (497.9) |
| Ponderal index mean (SD) | - | - | 25.7 (4.1) | 25.8 (4.2) | 25.4 (3.3) |

*those with MRI vs those without $p<0.01$; Abbreviations: SES socioeconomic status; Father's SES is based on five categories from professional (1) to unskilled labour; (higher number indicates lower SES); - is used when data were not available.

**Supplementary Table 7: Multiple regression analysis of early life factors and cSVD in STRADL, the Dutch Famine Birth Cohort, the LBC 1936 and the Simpson's cohort.**

| | STRADL | | Dutch Famine | | LBC | | Simpson's | | Meta-analysis | |
|---|---|---|---|---|---|---|---|---|---|---|
| | OR (95% CI)[a] | p | OR (95% CI)[b] | p | OR (95% CI)[b] | p | OR (95% CI)[b] | p | OR (95% CI) | p |
| **Moderate/severe cSVD** | | | | | | | | | | |
| Birth weight | 0·94 (0·86-1.04) | 0·22 | 0·95 (0·86-1.05) | 0·29 | 1·001 (0·92-1.10) | 0·98 | 0·94 (0·85-1·04) | 0·21 | 0·96 (0·92-1·01) | 0·08 |
| Low education | 1·41 (0·41-4·78) | 0·59 | 0·72 (0·27-1.95) | 0·52 | 1·23 (0·31-4·85) | 0·77 | 1·08 (0·34-3·40) | 0·90 | 1·02 (0·57-1·82) | 0·95 |
| Manual Father's occupation | 1·62 (0·34-7·72) | 0·54 | 0·67 (0·23-1.91) | 0·45 | 0·55 (0·19-1·58) | 0·26 | 0·40 (0·16-1·01) | 0·05 | 0·59 (0·34-1·02) | 0·06 |
| **Moderate/severe WMH** | | | | | | | | | | |
| Birth weight | 0·93 (0·83-1·04) | 0·21 | 1·02 (0·93-1·12) | 0·68 | 0·99 (0·90-1·08) | 0·79 | **0·90 (0·81-0·999)** | **0·048** | 0·96 (0·91-1·02) | 0·20 |
| Low education | 1·12 (0·22-5·82) | 0·89 | 0·53 (0·21-1·37) | 0·19 | 0·81 (0·29-2·84) | 0·74 | 0·88 (0·28-2·7) | 0·82 | 0·74 (0·42-1·28) | 0·28 |
| Manual Father's occupation | na | na | 1.29 (0·47-3·58) | 0·63 | 1·03 (0·33-3·21) | 0·96 | 0·85 (0·33-2·22) | 0·75 | 1·03 (0·57-1·86) | 0·92 |
| **Presence of 1+ lacune** | | | | | | | | | | |
| Birth weight | 0·91 (0·83-1.01) | 0·07 | 0·94 (0·85-1·03) | 0·17 | 0·99 (0·86-1·14) | 0·89 | 0·95 (0·86-1·05) | 0·31 | **0·94 (0·89-0·99)** | **0·02** |
| Low education | 0·64 (0·13-3·04) | 0·57 | 1·45 (0·53-3·95) | 0·47 | 0·86 (0·09-8·13) | 0·89 | 1·11 (0·34-3·59) | 0·86 | 1·11 (0·57-2·15) | 0·76 |
| Manual Father's occupation | 0·84 (0·21-3·32) | 0·80 | 0·86 (0·30-2·41) | 0·86 | 0·34 (0·07-1·58) | 0·17 | 0·42 (0·17-1·05) | 0·07 | 0·58 (0·32-1·04) | 0·07 |
| **Presence of 1+ CMB** | | | | | | | | | | |
| Birth weight | 0·97 (0·87-1·08) | 0·55 | 0·92 (0·81-1·05) | 0·20 | 1·06 (0·95-1·18) | 0·33 | 1·08 (0·93-1·26) | 0·32 | 1·00 (0·93-1·08) | 0·94 |
| Low education | 0·69 (0·14-3·30) | 0·64 | 1·05 (0·30-3·74) | 0·94 | 1·06 (0·21-5·45) | 0·94 | 4·07 (0·43-38·82) | 0·22 | 1·12 (0·51-2·47) | 0·77 |
| Manual Father's occupation | 1·38 (0·28-6·72) | 0·69 | 0·39 (0·11-1·44) | 0·16 | 0·42 (0·13-1·42) | 0·17 | 0·44 (0·12-1·65) | 0·22 | **0·51 (0·26-0·98)** | **0·04** |
| **Presence of 1+ infarct** | | | | | | | | | | |
| Birth weight | 0·91 (0·81-1·02) | 0·09 | 0·98 (0·89-1·09) | 0·72 | 0·94 (0·85-1·04) | 0·26 | 0·93 (0·80-1·08) | 0·35 | **0·94 (0·89-1·00)** | **0·03** |
| Low education | 0·43 (0·05-3·53) | 0·43 | 1·86 (0·62-5·63) | 0·27 | 2·91 (0·35-24·49) | 0·33 | 0·92 (0·17-4·84) | 0·92 | 1·40 (0·64-3·07) | 0·40 |
| Manual Father's occupation | 0·54 (0·13-2·30) | 0·40 | 1·74 (0·54-5·66) | 0·36 | 0·49 (0·16-1·54) | 0·22 | 0·37 (0·10-1·40) | 0·37 | 0·66 (0·33-1·33) | 0·25 |
| **Moderate/severe atrophy** | | | | | | | | | | |
| Birth weight | 0·89 (0·78-1·01) | 0·07 | 1·01 (0·92-1·12) | 0·80 | 0·97 (0·90-1·05) | 0·47 | 1·01 (0·91-1·12) | 0·87 | 0·97 (0·93-1·02) | 0·31 |
| Low education | 0·71 (0·08-6·25) | 0·75 | 1·46 (0·52-4·12) | 0·48 | 1·35 (0·44-4·12) | 0·60 | 0·51 (0·17-1·48) | 0·22 | 0·99 (0·54-1·80) | 0·97 |
| Manual Father's occupation | 1·15 (0·12-10·61) | 0·90 | 0·94 (0·32-2·75) | 0·91 | 0·82 (0·31-2·16) | 0·82 | 0·67 (0·26-1·73) | 0·41 | 0·76 (0·48-1·21) | 0·25 |
| **Moderate/severe EPVS** | | | | | | | | | | |
| Birth weight | **0·92 (0·86-0·98)** | **0·01** | 0·95 (0·87-1·04) | 0·29 | 0·97 (0·90-1·05) | 0·47 | 0·98 (0·89-1·08) | 0·68 | **0·95 (0·91-0·99)** | **0·007** |

| | | | | | | | | | | |
|---|---|---|---|---|---|---|---|---|---|---|
| Low education | 0·97 (0·42-2·27) | 0·94 | 0·63 (0·24-1.64) | 0·34 | 2·40 (0·68-8·43) | 0·17 | 1.36 (0·45-4·14) | 0·59 | 1.07 (0·65-1.77) | 0·80 |
| Manual Father's occupation | 0·70 (0·31-1.60) | 0·40 | 0·99 (0·35-2·80) | 0·99 | 0·53 (0·20-1.39) | 0·53 | 0·54 (0·20-1.51) | 0·24 | 0·66 (0·41-1.06) | 0·09 |

cSVD= cerebral small vessel disease; WMH= white matter hyperintensities; CMB= cerebral microbleed; EPVS= enlarged perivascular spaces
NA: due to the low cSVD burden in STRADL participants it was not possible to conduct all analyses.

[a] Analyses adjusted for variables in the table; [b] adjusted for variables in the table and age and sex

#### Additional results of associations between early life factors and SVD.

##### Ponderal index and SVD.

Ponderal index was not associated with SVD in the meta-analysis but point estimates were in the expected direction (higher ponderal index associated with fewer lesions) for total SVD burden, WMH burden, lacunes, infarcts and PVS.

Increasing ponderal index was not associated with WMH volume or brain volume in the Dutch Famine Birth Cohort, the LBC1936 or Simpson's cohort (Supplementary Table 4).

#### Results of associations between early life factors and SVD in individual cohorts.

##### Ponderal index

In the LBC1936, increasing ponderal index was associated with decreased risk of total SVD (per kg/m3 OR=0.85 95%CI=0.74-0.98, Supplementary Fig. 2) independent of age, sex, hypertension, smoking behaviour and adult SES.

Increasing ponderal index was associated with increased risk of cerebral micro-bleeds in the Simpson's cohort but not the expected direction (per kg/m3 OR=1.25 95%CI=1.03-1.50, Supplementary Fig. 2).

##### Childhood SES

In individual cohorts, low childhood SES was associated with lower total SVD burden (Simpson's cohort only OR=0.36, 95%CI=0.14-0.93) and decreased risk of infarcts (STRADL OR=0.52, 95%CI=0.28-0.94) none of which were in the expected direction (Figure 1D).

**Supplementary Fig. 2: Forest plot showing associations between features of SVD and ponderal index in the Dutch Famine Birth cohort, LBC 1936 and Simpson's cohort . All analyses are adjusted for age, sex, hypertension, smoking behaviour, adult SES and gestational age. cSVD = total SVD burden**

| Study or Subgroup | log[Odds Ratio] | SE | Weight | Odds Ratio IV, Random, 95% CI |
|---|---|---|---|---|
| **1.4.1 Moderate/severe total cSVD** | | | | |
| Dutch Famine Birth Cohort | -0.0534 | 0.1141 | 17.3% | 0.95 [0.76, 1.19] |
| LBC 1936 | -0.1625 | 0.0707 | 36.3% | 0.85 [0.74, 0.98] |
| Simpson Cohort | -0.0101 | 0.0584 | 46.4% | 0.99 [0.88, 1.11] |
| **Subtotal (95% CI)** | | | **100.0%** | **0.93 [0.84, 1.03]** |
| Heterogeneity: Tau² = 0.00; Chi² = 2.78, df = 2 (P = 0.25); I² = 28% | | | | |
| Test for overall effect: Z = 1.42 (P = 0.16) | | | | |
| **1.4.2 Moderate/severe WMH** | | | | |
| Dutch Famine Birth Cohort | 0.1044 | 0.1127 | 19.2% | 1.11 [0.89, 1.38] |
| LBC 1936 | -0.0943 | 0.0657 | 41.0% | 0.91 [0.80, 1.04] |
| Simpson Cohort | -0.1165 | 0.0673 | 39.8% | 0.89 [0.78, 1.02] |
| **Subtotal (95% CI)** | | | **100.0%** | **0.94 [0.84, 1.04]** |
| Heterogeneity: Tau² = 0.00; Chi² = 3.00, df = 2 (P = 0.22); I² = 33% | | | | |
| Test for overall effect: Z = 1.18 (P = 0.24) | | | | |
| **1.4.3 Presence of 1+ lacune** | | | | |
| Dutch Famine Birth Cohort | -0.1347 | 0.1118 | 20.1% | 0.87 [0.70, 1.09] |
| LBC 1936 | -0.1625 | 0.1369 | 13.4% | 0.85 [0.65, 1.11] |
| Simpson Cohort | -0.0576 | 0.0614 | 66.5% | 0.94 [0.84, 1.06] |
| **Subtotal (95% CI)** | | | **100.0%** | **0.92 [0.83, 1.01]** |
| Heterogeneity: Tau² = 0.00; Chi² = 0.72, df = 2 (P = 0.70); I² = 0% | | | | |
| Test for overall effect: Z = 1.74 (P = 0.08) | | | | |
| **1.4.4 Presence of 1+ CMBs** | | | | |
| Dutch Famine Birth Cohort | 0.1062 | 0.1353 | 19.3% | 1.11 [0.85, 1.45] |
| LBC 1936 | 0.0198 | 0.0696 | 47.9% | 1.02 [0.89, 1.17] |
| Simpson Cohort | 0.2207 | 0.0951 | 32.8% | 1.25 [1.03, 1.50] |
| **Subtotal (95% CI)** | | | **100.0%** | **1.11 [0.97, 1.26]** |
| Heterogeneity: Tau² = 0.00; Chi² = 2.92, df = 2 (P = 0.23); I² = 31% | | | | |
| Test for overall effect: Z = 1.55 (P = 0.12) | | | | |
| **1.4.5 Presence of 1+ infarct** | | | | |
| Dutch Famine Birth Cohort | -0.0101 | 0.1216 | 23.9% | 0.99 [0.78, 1.26] |
| LBC 1936 | -0.1278 | 0.0884 | 45.2% | 0.88 [0.74, 1.05] |
| Simpson Cohort | -0.1054 | 0.1068 | 31.0% | 0.90 [0.73, 1.11] |
| **Subtotal (95% CI)** | | | **100.0%** | **0.91 [0.81, 1.02]** |
| Heterogeneity: Tau² = 0.00; Chi² = 0.63, df = 2 (P = 0.73); I² = 0% | | | | |
| Test for overall effect: Z = 1.56 (P = 0.12) | | | | |
| **1.4.6 Moderate/severe atrophy** | | | | |
| Dutch Famine Birth Cohort | 0.0392 | 0.1275 | 9.4% | 1.04 [0.81, 1.34] |
| LBC 1936 | 0.0392 | 0.0516 | 57.5% | 1.04 [0.94, 1.15] |
| Simpson Cohort | -0.0408 | 0.0681 | 33.0% | 0.96 [0.84, 1.10] |
| **Subtotal (95% CI)** | | | **100.0%** | **1.01 [0.94, 1.09]** |
| Heterogeneity: Tau² = 0.00; Chi² = 0.92, df = 2 (P = 0.63); I² = 0% | | | | |
| Test for overall effect: Z = 0.33 (P = 0.74) | | | | |
| **1.4.7 Moderate/severe EPVS** | | | | |
| Dutch Famine Birth Cohort | -0.102 | 0.1134 | 12.4% | 0.90 [0.72, 1.13] |
| LBC 1936 | -0.1098 | 0.0578 | 47.9% | 0.90 [0.80, 1.00] |
| Simpson Cohort | -0.009 | 0.0635 | 39.7% | 0.99 [0.88, 1.12] |
| **Subtotal (95% CI)** | | | **100.0%** | **0.93 [0.86, 1.01]** |
| Heterogeneity: Tau² = 0.00; Chi² = 1.48, df = 2 (P = 0.48); I² = 0% | | | | |
| Test for overall effect: Z = 1.72 (P = 0.09) | | | | |

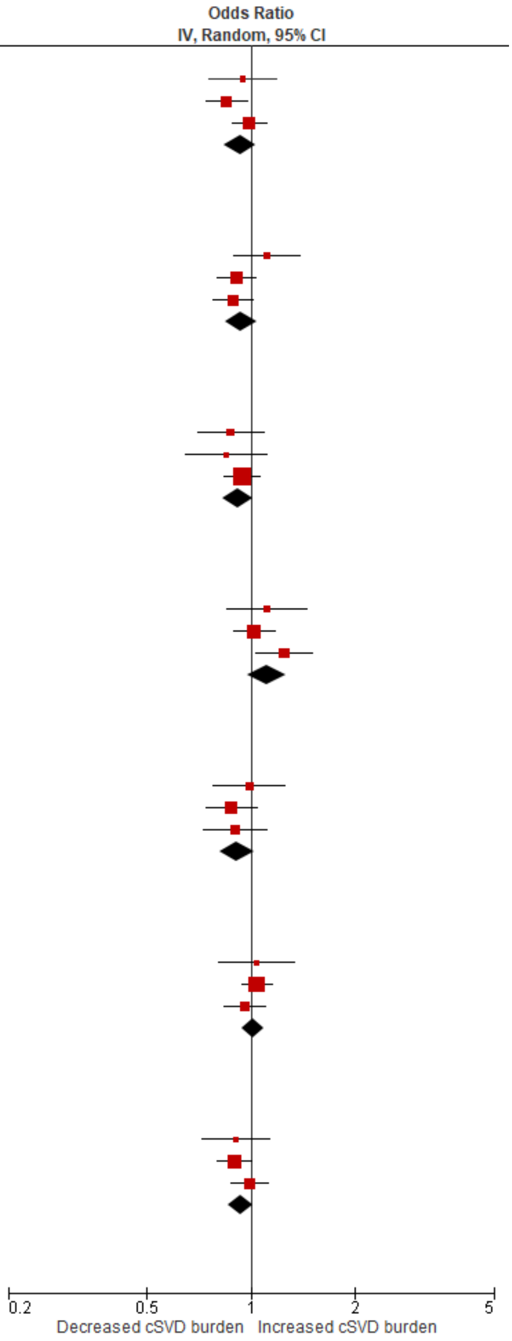


Test for subgroup differences: Chi² = 9.27, df = 6 (P = 0.16), I² = 35.2%

**Supplementary Table 8: Associations between early life factors and whole brain volume as a percentage of the ICV and WMH volume in STRADL, the LBC1936 and Simpson's cohort. All analysis are adjusted for age, sex, hypertension, smoking behaviour, adult SES**

| | **STRADL** | | **Dutch Famine Birth cohort** | | **LBC1936** | | **Simpson's** | | **Meta-analysis** | |
|---|---|---|---|---|---|---|---|---|---|---|
| | B (95% CI) | p | B (95% CI) | p | B (95% CI) | p | B (95% CI) | p | B (95% CI) | p |
| **Whole brain volume normalised for ICV** | | | | | | | | | | |
| Ponderal index* | - | - | - | - | 0·09 (-0·001-0·001) | 0·41 | -0·14 (-0·09-0·02) | 0·29 | -0·04 (-0·18-0·11) | 0·39 |
| Birth weight (per 100g)* | 0·11 (-0·001-0·004) | 0·16 | - | - | 0·04 (-0·13-0·21) | 0·62 | -0·20 (-0·39-0·01) | 0·40 | -0·06 (-0·22-0·10) | 0·45 |
| Childhood IQ | -0·06 (-0·002-0·001) | 0·34 | - | - | 0·04 (-0·04-0·12) | 0·30 | -0·09 (-0·55-0·41)** | 0·73 | 0·04 (-0·04-0·12) | 0·31 |
| **WMH volume normalised for ICV** | | | | | | | | | | |
| Ponderal index* | - | - | 0·10 (-0·14-0·59) | 0·27 | -0·17 (-0·38-0·06) | 0·11 | -0·11 (-0·30-0·09) | 0·19 | -0·05 (-0·18-0·09) | 0·48 |
| Birth weight (per 100g)* | - | - | 0·07 (-0·13-0·27) | 0·38 | -0·08 (-0·25-0·09) | 0·40 | -0·08 (-0·27-0·11) | 0·43 | -0·03 (-0·14-0·07) | 0·54 |
| Childhood IQ | -0·03 (-0·07-0·05) | 0·44 | - | - | -0·07 (-0·15-0·01) | 0·12 | 0·14 (-0·35-0·57)** | 0·30 | -0·06 (-0·14-0·01) | 0·11 |

* This analysis is also adjusted for gestational age; ** due to sample size this analysis is only adjusted for age and sex; - data were not available

#### Supplementary Table 9: Multiple regression analysis of Childhood IQ, education, childhood SES and cSVD in STRADL and the LBC 1936

NOTE: It was not possible to include the Simpson's cohort in this analysis due to the small number of participants with childhood IQ scores. Childhood IQ was not available in the Dutch Famine Birth cohort therefore this cohort was not included in this analysis.

All analyses are adjusted for each early life factor in the table and age, sex, hypertension, smoking behaviour and adult SES.

| | STRADL | | LBC 1936 | | Meta-analysis | |
|---|---|---|---|---|---|---|
| | OR (95% CI) | p | OR (95% CI) | p | OR (95% CI) | p |
| **Moderate/severe cSVD** | | | | | | |
| Childhood IQ | 0·97 (0·93-1.004) | 0·06 | 0·99 (0·97-1.00) | 0·07 | **0·98 (0·97-0·997)** | **0·02** |
| Low education | 0·87 (0·42-1.80) | 0·71 | 0·99 (0·56-1.75) | 0·97 | 0·94 (0·60-1.48) | 0·80 |
| Manual Father's occupation | 1.11 (0·54-2·28) | 0·78 | 1.32 (0·75-2·32) | 0·34 | 1.24 (0·79-1.93) | 0·35 |
| **Moderate/severe WMH**** | | | | | | |
| Childhood IQ* | 0·98 (0·94-1.02) | 0·34 | 0·99 (0·98-1.01) | 0·29 | 0·99 (0·98-1.00) | 0·20 |
| Low education | 1.19 (0·46-3·06) | 0·72 | 0·79 (0·47-1.33) | 0·37 | 0·87 (0·55-1.37) | 0·54 |
| Manual Father's occupation | 1.28 (0·51-3·21) | 0·60 | 1.25 (0·75-2·08) | 0·39 | 1.26 (0·80-1.96) | 0·32 |
| **Presence of 1+ lacune** | | | | | | |
| Childhood IQ* | 1.02 (0·95-1.09) | 0·66 | 0·97 (0·95-1.00) | 0·04 | 0·98 (0·95-1.02) | 0·29 |
| Low education | 0·34 (0·10-1.16) | 0·08 | 1.10 (0·38-3·18) | 0·86 | 0·64 (0·20-2·01) | 0·44 |
| Manual Father's occupation | 2·74 (0·79-9·50) | 0·11 | **0·30 (0·13-0·71)** | **0·01** | 0·87 (0·10-7·48) | 0·90 |
| **Presence of 1+ CMB** | | | | | | |
| Childhood IQ* | **0·95 (0·90-0·995)** | **0·03** | 0·995 (0·98-1.01) | 0·59 | 0·98 (0·93-1.02) | 0·32 |
| Low education | 1.06 (0·42-2·67) | 0·91 | 1.35 (0·67-2·71) | 0·41 | 1.24 (0·71-2·18) | 0·45 |
| Manual Father's occupation | 0·78 (0·33-1.87) | 0·58 | 1.20 (0·62-2·31) | 0·58 | 1.02 (0·61-1.73) | 0·93 |
| **Presence of 1+ infarct** | | | | | | |
| Childhood IQ* | 0·98 (0·89-1.08) | 0·68 | **0·98 (0·97-1.00)** | **0·03** | **0·98 (0·97-0·997)** | **0·02** |
| Low education | 1.14 (0·20-6·50) | 0·88 | 1.07 (0·56-2·05) | 0·84 | 0·95 (0·60-1.48) | 0·80 |
| Manual Father's occupation | 0·67 (0·13-3·45) | 0·63 | 0·77 (0·43-1.38) | 0·38 | 1.24 (0·79-1.93) | 0·35 |
| **Moderate/severe atrophy** | | | | | | |
| Childhood IQ* | 1.04 (0·98-1.10) | 0·24 | 0·98 (0·99-1.01) | 0·67 | 1.01 (0·96-1.05) | 0·79 |
| Low education | 0·60 (0·19-1.87) | 0·37 | 0·89 (0·57-1.35) | 0·58 | 0·82 (0·55-1.23) | 0·34 |

| | | | | | | |
|---|---|---|---|---|---|---|
| Manual Father's occupation | 2·12 (0·67-6·68) | 0·20 | 0·88 (0·59-1.31) | 0·51 | 1.13 (0·48-2·64) | 0·78 |
| **Moderate/severe EPVS** | | | | | | |
| Childhood IQ* | 0·98 (0·94-1.02) | 0·37 | 1.01 (0·99-1.02) | 0·48 | 1.00 (0·98-1.02) | 0·85 |
| Low education | 1.29 (0·58-2·90) | 0·53 | 0·91 (0·59-1.38) | 0·65 | 0·99 (0·67-1.44) | 0·94 |
| Manual Father's occupation | 0·82 (0·38-1.78) | 0·62 | 1.04 (0·69-1.55) | 0·86 | 0·99 (0·69-1.42) | 0·94 |

* OR is given per 1 point increase in IQ; moderate/severe WMH= Fazekas deep WMH score ≥2 or periventricular WMH score

**Supplementary Fig. 3: Scatterplots of association between age 11 IQ and a) WMH volume b) whole brain volume in STRADL, the LBC 1936 and Simpson's cohort.**

a)

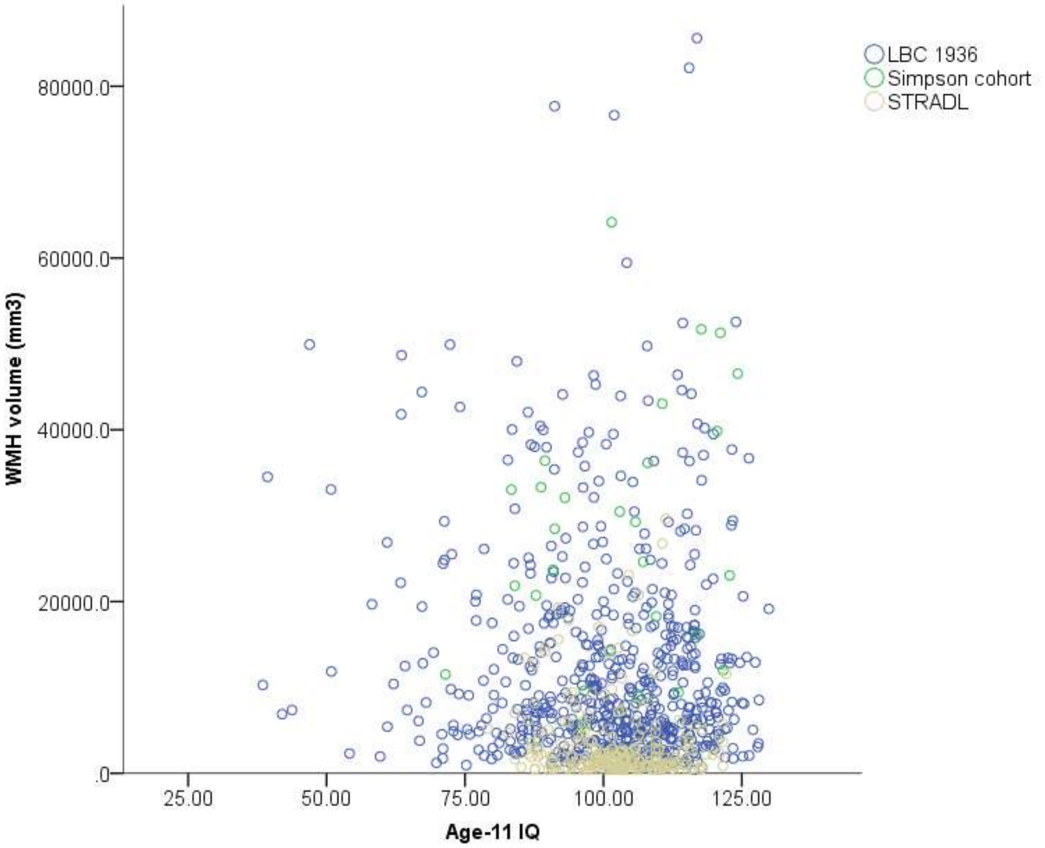

b)

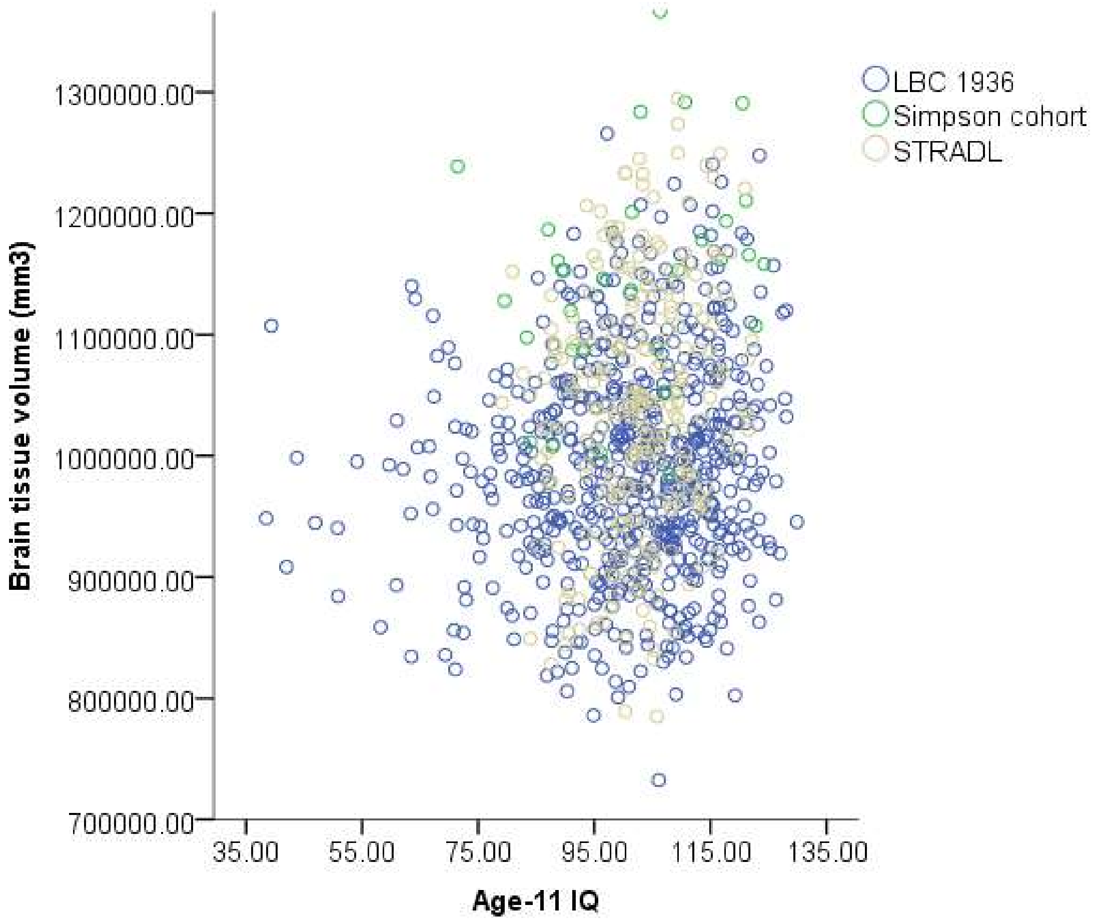

LBC 1936
Simpson cohort
STRADL
Brain tissue volume (mm3)
1300000.00
1200000.00
1100000.00
1000000.00
900000.00
800000.00
700000.00
35.00
55.00
75.00
95.00
115.00
135.00
Age-11 IQ

**Supplementary Fig. 4: Forest plots showing mean white matter hyperintensity (WMH) volume for those with a) low vs high levels of education b) low vs high childhood SES, random effects model for the mean difference.**

**a)**

| Study or Subgroup | Low education Mean | Low education SD | Low education Total | High education Mean | High education SD | High education Total | Weight | Mean Difference IV, Random, 95% CI |
|---|---|---|---|---|---|---|---|---|
| STRADL | 369.3054 | 446.2846 | 112 | 284.6279 | 488.4692 | 358 | 71.3% | 84.68 [-12.23, 181.59] |
| Dutch Famine Birth Cohort | 404.32466 | 430.843009 | 73 | 476.86977 | 625.637723 | 43 | 15.0% | -72.55 [-284.05, 138.96] |
| LBC 1936 | 1,186.1671 | 1,129.7139 | 432 | 1,234.3786 | 1,297.4641 | 168 | 13.4% | -48.21 [-271.46, 175.04] |
| Simpson Cohort | 3,101.1835 | 2,586.057 | 87 | 3,193.9707 | 3,017.6053 | 20 | 0.3% | -92.79 [-1522.58, 1337.00] |
| **Total (95% CI)** | | | **704** | | | **589** | **100.0%** | **42.72 [-39.10, 124.54]** |

Heterogeneity: Tau² = 0.00; Chi² = 2.53, df = 3 (P = 0.47); I² = 0%
Test for overall effect: Z = 1.02 (P = 0.31)

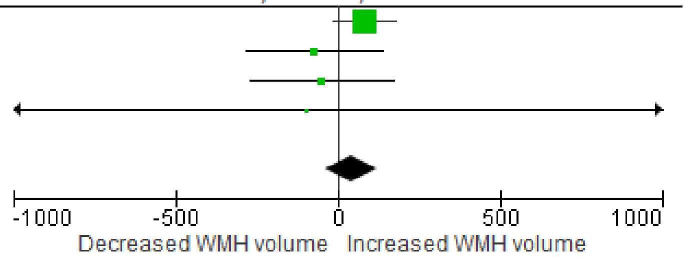


Negative mean difference= low education (<11 years) decreases WMH volume; positive difference= low education increases WMH volume.

**b)**

| Study or Subgroup | Low SES Mean | Low SES SD | Low SES Total | High SES Mean | High SES SD | High SES Total | Weight | Mean Difference IV, Random, 95% CI |
|---|---|---|---|---|---|---|---|---|
| STRADL | 320.486 | 433.0174 | 308 | 249.6962 | 446.8204 | 157 | 64.4% | 70.79 [-14.20, 155.78] |
| Dutch Famine Birth Cohort | 433.295 | 440.784095 | 62 | 523.97813 | 446.8204 | 32 | 19.5% | -90.68 [-280.43, 99.07] |
| LBC 1936 | 1,217.5348 | 1,185.047 | 152 | 1,147.0013 | 1,159.965 | 448 | 15.4% | 70.53 [-146.33, 287.39] |
| Simpson Cohort | 2,951.0752 | 2,592.6549 | 74 | 3,494.0246 | 2,799.9478 | 33 | 0.6% | -542.95 [-1666.14, 580.24] |
| **Total (95% CI)** | | | **596** | | | **670** | **100.0%** | **35.31 [-54.49, 125.12]** |

Heterogeneity: Tau² = 1378.52; Chi² = 3.42, df = 3 (P = 0.33); I² = 12%
Test for overall effect: Z = 0.77 (P = 0.44)

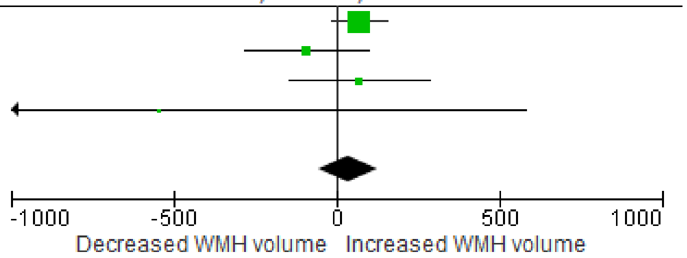


Negative mean difference= low SES (manual paternal occupation) decreases WMH volume; positive difference= low SES increases WMH volume.

**Supplementary Fig. 5: Forest plots showing total brain volume for those with a) low vs high levels of education b) low vs high childhood SES, random effects model for the mean difference.**

**a)**

| Study or Subgroup | Low education Mean | SD | Total | High education Mean | SD | Total | Weight | Mean Difference IV, Random, 95% CI |
|---|---|---|---|---|---|---|---|---|
| STRADL | 10,617.6675 | 1,022.9806 | 248 | 10,728.7594 | 1,110.6724 | 798 | 47.1% | -111.09 [-259.91, 37.73] |
| LBC 1936 | 9,870.8599 | 889.4543 | 432 | 10,051.481 | 869.1933 | 168 | 45.0% | -180.62 [-336.54, -24.70] |
| Simpson's Cohort | 11,250.4818 | 949.4492 | 87 | 11,824.3877 | 1,040.7666 | 20 | 7.9% | -573.91 [-1071.76, -76.05] |
| **Total (95% CI)** | | | **767** | | | **986** | **100.0%** | **-178.86 [-325.07, -32.66]** |

Heterogeneity: Tau² = 6040.58; Chi² = 3.13, df = 2 (P = 0.21); I² = 36%
Test for overall effect: Z = 2.40 (P = 0.02)

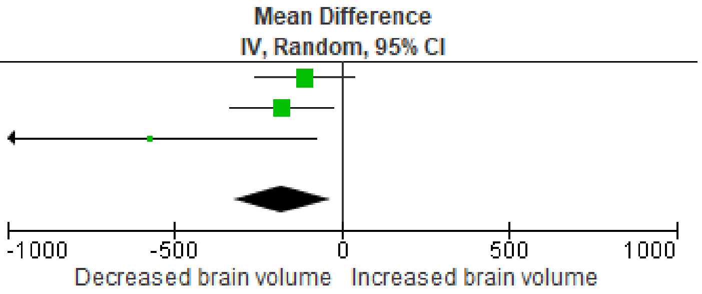


Negative mean difference= low education (<11 years) decreases brain volume volume; positive difference= low education increases brain volume

**b)**

| Study or Subgroup | Low SES Mean | SD | Total | High SES Mean | SD | Total | Weight | Mean Difference IV, Random, 95% CI |
|---|---|---|---|---|---|---|---|---|
| STRADL | 10,600.4456 | 1,090.3803 | 701 | 10,909 | 1,064.2719 | 345 | 52.1% | -308.55 [-446.86, -170.25] |
| LBC 1936 | 9,904.5153 | 899.5036 | 152 | 9,971.299 | 849.2843 | 448 | 47.9% | -66.78 [-229.98, 96.41] |
| Simpson's Cohort | 11,307.463 | 96,551.9905 | 74 | 11,471.462 | 1,042.7152 | 33 | 0.0% | -164.00 [-22165.40, 21837.40] |
| **Total (95% CI)** | | | **927** | | | **826** | **100.0%** | **-192.74 [-403.96, 18.47]** |

Heterogeneity: Tau² = 17313.15; Chi² = 4.91, df = 2 (P = 0.09); I² = 59%
Test for overall effect: Z = 1.79 (P = 0.07)

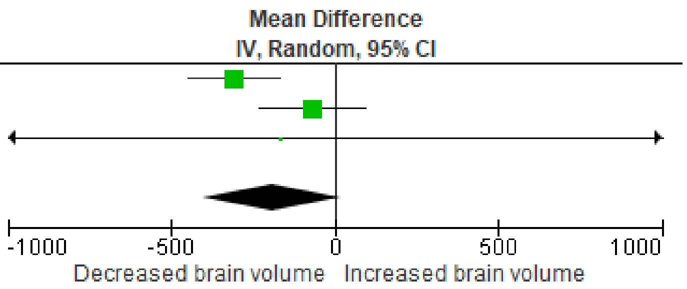


Negative mean difference= low SES (manual paternal occupation) decreases brain volume; positive difference= low SES increases brain volume.

**Supplementary Table 10: Associations between early life factors and whole brain volume as a percentage of the ICV and WMH volume in STRADL, the LBC 1936 and Simpson's cohort. All analysis are adjusted for age, sex, hypertension, smoking behaviour, adult SES.**

| | **STRADL** | | **Dutch Famine Birth cohort** | | **LBC 1936** | | **Simpson's** | | **Meta-analysis** | |
|---|---|---|---|---|---|---|---|---|---|---|
| | B (95% CI) | p | B (95% CI) | p | B (95% CI) | p | B (95% CI) | p | B (95% CI) | p |
| **Whole brain volume as a % of ICV** | | | | | | | | | | |
| Low vs high education level | 0·01 (-0·01-0·02) | 0·76 | - | - | 0·03 (-0·05-0·11) | 0·46 | -0·14 (-0·06-0·33) | 0·22 | 0·01(-0·04-0·06) | 0·75 |
| Low vs high childhood SES | -0·01 (-0·02-0·01) | 0·81 | - | - | 0·04 (-0·04-0·12) | 0·14 | -0·09 (-0·29-0·11) | 0·40 | 0·01(-0·05-0·07) | 0·74 |
| **WMH volume** | | | | | | | | | | |
| Low vs high education level | 0·06 (0·00-0·001) | 0·21 | 0·08 (-0·12-0·28) | 0·38 | 0·004 (-0·07-0·08) | 0·90 | -0·01 (-0·21-0·19) | 0·97 | 0·02 (-0·03-0·08) | 0·33 |
| Low vs high childhood SES | -0·02 (-0·001-0·001) | 0·61 | 0·16 (-0·10-0·50) | 0·28 | 0·01 (-0·07-0·09) | 0·85 | -0·17 (-0·36-0·03) | 0·08 | -0·01 (-0·06-0·51) | 0·87 |

**Supplementary tables 8a-d**: **Univariate logistic regressions for associations between early life factors and markers of SVD.**

**a) STRADL**

| | Total SVD | WMH | EPVS | Lacunes | CMBs | Infarcts | Atrophy |
|---|---|---|---|---|---|---|---|
| | OR (95%CI) | | | | | | |
| Age | **1.12 (1.09-1.14)** | **1.11 (1.08-1.15)** | **1.09 (1.08-1.11)** | **1.11 (1.08-1.15)** | **1.08 (1.05-1.11)** | **1.04 (1.01-1.07)** | **1.06 (1.03-1.09)** |
| Sex | 0.84 (0.61-1.17) | 1.28 (0.86-1.93) | **0.73 (0.57-0.93)** | 0.81 (0.52-1.27) | 0.68 (0.47-1.01) | 0.62 (0.37-1.05) | **0.26 (0.15-0.44)** |
| Hypertension | **2.54 (1.82-3.53)** | **2.05 (1.38-3.06)** | **2.51 (1.90-3.33)** | 1.43 (0.90-2.28) | **2.66 (1.80-3.93)** | 1.43 (0.82-2.47) | **1.69 (1.03-2.79)** |
| Ever smoker | 1.23 (0.88-1.72) | 1.41 (0.94-2.11) | 1.01 (0.78-1.30) | 0.76 (0.47-1.22) | 1.27 (0.85-1.92) | 1.44 (0.82-2.52) | **1.98 (1.17-3.33)** |
| Manual adult SES | 1.16 (0.83-1.62) | 1.27 (0.85-1.91) | **0.75 (0.57-0.97)** | 1.44 (0.92-2.27) | 1.02 (0.68-1.54) | 1.20 (0.69-2.09) | 0.80 (0.47-1.39) |
| Birth weight | 0.94 (0.85-1.03) | 0.91 (0.81-1.03) | 0.93 (0.87-0.99) | 0.91 (0.83-1.01) | 0.97 (0.87-1.08) | 0.92 (0.82-1.02) | 0.88 (0.78-1.01) |
| Childhood IQ | 0.98 (0.95-1.01) | 0.98 (0.94-1.01) | 0.98 (0.95-1.02) | 0.99 (0.94-1.04) | 0.98 (0.94-1.03) | 0.98 (0.91-1.06) | 1.01 (0.97-1.06) |
| Education | **1.58 (1.11-2.24)** | 1.36 (0.89-2.10) | 1.17 (0.88-1.55) | 1.34 (0.82-2.19) | **1.68 (1.11-2.54)** | 1.07 (0.59-1.95) | 0.88 (0.49-1.59) |
| Manual childhood SES | 0.87 (0.62-1.21) | 1.16 (0.76-1.77) | 0.95 (0.74-1.23) | 0.97 (0.61-1.54) | 0.78 (0.53-1.17) | 0.72 (0.43-1.23) | 1.06 (0.63-1.78) |

**b) Dutch Famine Birth cohort**

| | Total SVD | WMH | EPVS | Lacunes | CMBs | Infarcts | Atrophy |
|---|---|---|---|---|---|---|---|
| OR (95% CI) | | | | | | | |
| Age | 0.96 (0.60-1.62) | 1.38 (0.87-2.21) | 1.31 (0.81-2.12) | 0.71 (0.42-1.20) | 0.69 (0.36-1.32) | 1.04 (0.61-1.80) | 1.15 (0.69-1.91) |

| | | | | | | | |
|---|---|---|---|---|---|---|---|
| Sex | **2.79 (1.07-7.23)** | 1.83 (0.77-4.34) | 1.92 (0.78-4.73) | 1.34 (0.55-3.27) | 0.99 (0.34-2.87) | 0.60 (0.23-1.51) | 0.83 (0.33-2.07) |
| Hypertension | 0.79 (0.33-1.88) | 0.85 (0.37-1.95) | 1.05 (0.45-2.47) | 0.70 (0.29-1.69) | 0.87 (0.30-2.50) | 0.77 (0.30-1.98) | 1.08 (0.43-2.74) |
| Ever smoker | 0.63 (0.26-1.50) | 0.54 (0.24-1.26) | 1.52 (0.62-3.74) | 1.27 (0.51-3.15) | 0.81 (0.28-2.35) | 0.91 (0.35-2.33) | 2.73 (0.94-7.98) |
| Manual adult SES | 0.90 (0.36-2.25) | 0.65 (0.27-1.58) | 1.55 (0.65-3.72) | 1.07 (0.44-2.61) | 0.75 (0.24-2.34) | 2.4 (0.94-6.15) | 1.38 (0.55-3.48) |
| Birth weight | 0.95 (0.87-1.03) | 1.01 (0.93-1.09) | 0.94 (0.87-1.03) | 0.95 (0.87-1.04) | 0.94 (0.85-1.05) | 0.98 (0.91-1.09) | 1.02 (0.93-1.12) |
| Low education | 0.89 (0.37-2.15) | 0.71 (0.31-1.65) | 0.65 (0.27-1.54) | 1.16 (0.47-2.88) | 1.38 (0.48-4.29) | 1.34 (0.50-3.61) | 1.14 (0.44-2.97) |
| Manual childhood SES | 0.65 (0.30-2.10) | 1.27 (0.48-3.32) | 1.11 (0.42-2.96) | 1.00 (0.38-2.66) | 0.46 (0.13-1.55) | 1.65 (0.54-5.05) | 1.00 (0.36-2.79) |

**c) LBC1936**

| | Total SVD | WMH | EPVS | Lacunes | CMBs | Infarcts | Atrophy |
|---|---|---|---|---|---|---|---|
| | OR (95% CI) | | | | | | |
| Age | **1.38 (1.05-1.80)** | 1.28 (0.99-1.63) | 1.07 (0.87-1.32) | 1.31 (0.80-2.13) | **1.50 (1.08-2.09)** | 0.90 (0.73-1.10) | **1.45 (1.08-1.95)** |
| Sex | **1.54 (1.05-2.26)** | **1.56 (1.09-2.23)** | 0.88 (0.65-1.19) | 0.93 (0.46-1.88) | 0.83 (0.51-1.33) | 0.75 (0.49-1.15) | **0.45 (0.33-0.61)** |
| Hypertension | **1.70 (1.15-2.51)** | **2.15 (1.49-3.12)** | **1.53 (1.13-2.09)** | **2.90 (1.33-6.33)** | 1.07 (0.67-1.71) | **2.24 (1.44-3.48)** | 1.17 (0.87-1.58) |
| Ever smoker | 0.89 (0.60-1.30) | 1.02 (0.71-1.46) | 0.94 (0.69-1.27) | 1.60 (0.77-3.31) | 0.57 (0.35-0.91) | 1.27 (0.83-1.94) | 1.28 (0.95-1.73) |
| Manual adult SES | **1.63 (1.04-2.53)** | 1.37 (0.89-2.1) | 1.43 (0.98-2.01) | 1.17 (0.49-2.78) | 1.29 (0.74-2.25) | **1.77 (1.09-2.87)** | **1.49 (1.02-2.17)** |

| | | | | | | | |
|---|---|---|---|---|---|---|---|
| Birth weight | 0.99 (0.91-1.07) | 0.97 (0.99-1.05) | 0.98 (0.92-1.05) | 1.01 (0.88-1.16) | 1.06 (0.96-1.18) | 0.95 (0.87-1.04) | 0.98 (0.91-1.05) |
| Childhood IQ | **0.99 (0.97-0.99)** | **0.99 (0.98-1.00)** | 1.00 (0.99-1.01) | 0.99 (0.96-1.01) | 0.99 (0.98-1.01) | **0.98 (0.97-0.99)** | **0.99 (0.98-1.00)** |
| Low education | 1.43 (0.87-2.34) | 1.16 (0.78-1.75) | 1.04 (0.74-1.46) | 1.06 (0.48-2.32) | 1.64 (0.92-2.92) | 1.43 (0.87-2.34) | 0.96 (0.69-1.33) |
| Manual childhood SES | 1.58 (0.96-2.63) | 1.37 (0.86-2.16) | 0.99 (0.69-1.42) | 0.56 (0.26-1.21) | 1.32 (0.73-2.36) | 0.93 (0.56-1.55) | 0.84 (0.59-1.21) |

**d) Simpson's cohort**

| | Total SVD | WMH | EPVS | Lacunes | CMBs | Infarcts | Atrophy |
|---|---|---|---|---|---|---|---|
| OR 95% CI | | | | | | | |
| Age | 1.03 (0.77-1.38) | 1.08 (0.79-1.46) | 1.00 (0.74-1.36) | 1.11 (0.81-1.52) | 1.19 (0.77-1.85) | 1.00 (0.64-1.56) | 1.18 (0.86-1.63) |
| Sex | 0.80 (0.31-2.08) | 0.65 (0.26-1.63) | 0.98 (0.38-2.53) | 0.98 (0.38-2.55) | 0.39 (0.11-1.40) | 1.04 (0.26-4.25) | **0.39 (0.16-0.97)** |
| Hypertension | 1.61 (0.68-3.80) | 1.48 (0.62-3.53) | 0.83 (0.35-1.97) | 1.45 (0.60-3.46) | 2.21 (0.60-8.11) | 4.72 (0.96-23.19) | **2.47 (1.00-6.1)** |
| Ever smoker | 0.91 (0.39-2.15) | 1.58 (0.65-3.86) | 1.15 (0.48-2.76) | 1.62 (0.66-4.0) | 1.59 (0.44-5.85) | 1.91 (0.47-7.74) | 2.25 (0.88-5.74) |
| Manual adult SES | 1.41 (0.59-3.33) | **2.57 (1.06-6.25)** | 0.58 (0.24-1.40) | 0.97 (0.40-2.35) | 1.67 (0.47-5.89) | 1.73 (0.47-6.3) | **2.88 (1.16-7.14)** |
| Birth weight | 0.95 (0.86-1.04) | 0.91 (0.83-1.01) | 0.98 (0.89-1.08) | 0.96 (0.87-1.05) | 1.09 (0.95-1.26) | 0.94 (0.82-1.09) | 1.02 (0.93-1.13) |
| Childhood IQ | 0.96 (0.87-1.05) | 1.01 (0.96-1.07) | 1.01 (0.96-1.07) | 0.99 (0.93-1.05) | - | 1.00 (0.91-1.11) | 1.07 (0.98-1.17) |
| Low education | 0.94 (0.32-2.80 | 0.77 (0.26-2.24) | 1.30 (0.45-3.76) | 0.99 (0.32-3.02) | 2.64 (0.32-22.06) | 0.85 (0.17-4.29) | 0.41 (0.15-1.14) |

| | | | | | | | |
|---|---|---|---|---|---|---|---|
| Manual childhood SES | 0.42 (0.17-1.04) | 0.86 (0.34-2.18) | 0.56 (0.20-1.55) | 0.43 (0.17-1.07) | 0.47 (0.13-1.67) | 0.40 (0.11-1.45) | 0.64 (0.26-1.61) |
